# Adverse Drug Events Across Autoimmune Rheumatic Diseases: A Nested, Encounter-Matched Case-Control Study

**DOI:** 10.64898/2026.05.19.26352957

**Authors:** Ashley Adanna Lewis, Cho-Yi Huang, James Cragun, Lauren Vuong, Anushka Irani, Christine Anastasiou, Selen Bozkurt, Macarius Donneyong, Shivani Garg, Cornelius Botha Groenewald, Michael Weisman, Titilola Falasinnu

## Abstract

**Background:** Polypharmacy is common in autoimmune rheumatic diseases (ARDs) and increases adverse drug events (ADEs), but comparative evidence across diseases is limited. We aimed to quantify ADE burden and identify medications associated with ADE risk across six ARDs, and to examine shared and disease-specific patterns across diseases.

**Methods:** We conducted a retrospective cohort study at a tertiary medical center (2010–2024). Adults with ankylosing spondylitis (AS), psoriatic arthritis (PsA), rheumatoid arthritis (RA), Sjögren’s disease (SjD), systemic lupus erythematosus (SLE), or systemic sclerosis (SSc) were identified using diagnostic codes. ADEs were ascertained using validated case definitions. Medications were mapped to Anatomical Therapeutic Chemical classes; active exposure was defined within 30 days before the index date. Polypharmacy was defined as ≥5 concurrent medications (minor 5–10; major >10). Within each ARD, nested case–control analyses matched on encounter type (1:4) were performed, and adjusted odds ratios (aORs) were estimated using conditional logistic regression.

**Findings:** Among 10,578 patients, 3,154 (29.8%) experienced at least one ADE. ADE burden varied across diseases, with the highest prevalence observed in SSc (35.9%). Polypharmacy was common (57.3% minor, 39.4% major) and medication burden was consistently higher in ADE cases across encounter types (eg, SLE outpatient median 12 vs 6; inpatient 20 vs 10; emergency 17 vs 8). Across ARDs, the strongest associations with ADEs were observed for supportive and symptom-directed therapies (acid suppressors, pain adjuncts, and sedative-hypnotic/psychotropic medications), whereas conventional disease-modifying antirheumatic drugs (DMARDs) showed weaker associations. Disease-specific signatures included gastrointestinal agents in SSc (metoclopramide aOR 12.32), antibiotics and respiratory agents in AS (ciprofloxacin aOR 13.71, fluticasone aOR 8.88).

**Interpretation:** ADEs affect nearly one-third of ARD patients and increase with medication burden. Risk concentrates in supportive and symptom-directed therapies rather than DMARDs, with both shared and disease-specific patterns. Optimizing prescribing, particularly for pain management and corticosteroid use, can reduce medication-related harm.

## INTRODUCTION

Autoimmune rheumatic diseases (ARDs), including ankylosing spondylitis (AS), psoriatic arthritis (PsA), rheumatoid arthritis (RA), Sjögren’s disease (SjD), systemic lupus erythematosus (SLE), and systemic sclerosis (SSc), affect more than 4.3 million individuals in the United States.^1^ Management of ARDs requires long-term and often complex pharmacotherapy, including antimalarials, corticosteroids, conventional and biologic disease-modifying antirheumatic drugs (DMARDs), and targeted synthetic agents.^2,3^ In addition, therapies for comorbid conditions further expand the medication burden, wrestling in a high prevalence of polypharmacy.^4–6^

Polypharmacy is pervasive in ARDs and is associated with adverse drug events (ADEs), medication nonadherence, and greater healthcare utilization.^7,8^ For example, approximately 68% of patients with RA and 70% of those with SLE are exposed to five or more medications, more than double the prevalence in the general adult population (∼30%).^9^ This burden reflects a self-reinforcing cycle: disease activity and comorbidities necessitate multiple therapies, while treatment-related complications introduce additional medications.^10^As the number of medications increases, so does the risk of drug–drug interactions and adverse reactions. In RA, 81% of patients experience at least one potential interaction, and nearly 18% of these are classified as severe.^11^ In SLE, ADEs account for up to 44% of hospitalizations, yet the specific drugs, combinations, and biological mechanisms driving these events remain incompletely characterized.^12^

Existing studies have largely focused on individual diseases, particularly RA and SLE, limiting cross-disease comparisons and a broader understanding of medication-related harm. To address this gap, we conducted a comprehensive, multi-disease analysis across six major ARDs. We aimed to quantify ADE burden and identify medications associated with ADE risk across ARDs using a matched case–control design. Leveraging this unified framework, we further examined shared and disease-specific patterns across diseases. We hypothesized that ADE burden would be substantial across all ARDs and increase with medication burden, with both shared risk patterns driven by symptomatic treatment and disease-specific patterns reflecting variation in clinical presentation.

## METHODS

### Study Design and Data Source

We conducted a retrospective cohort study with a nested case–control analysis within each ARD cohort. For the case–control analysis, cases were defined as patients experiencing an ADE, and controls were matched on the same encounter types and ARD cohort but without ADEs. Figure 1 shows the study timeline and analytic windows. Electronic health record (EHR) data were extracted from Stanford Medicine’s STARR (Stanford Translational Research Integrated Database Technology) system. The study period spanned January 1, 2010, through December 31, 2024. We compared patients with and without ADEs to evaluate differences in medication burden, polypharmacy patterns, and drug exposures across ARDs and care settings.

**Figure 1.**
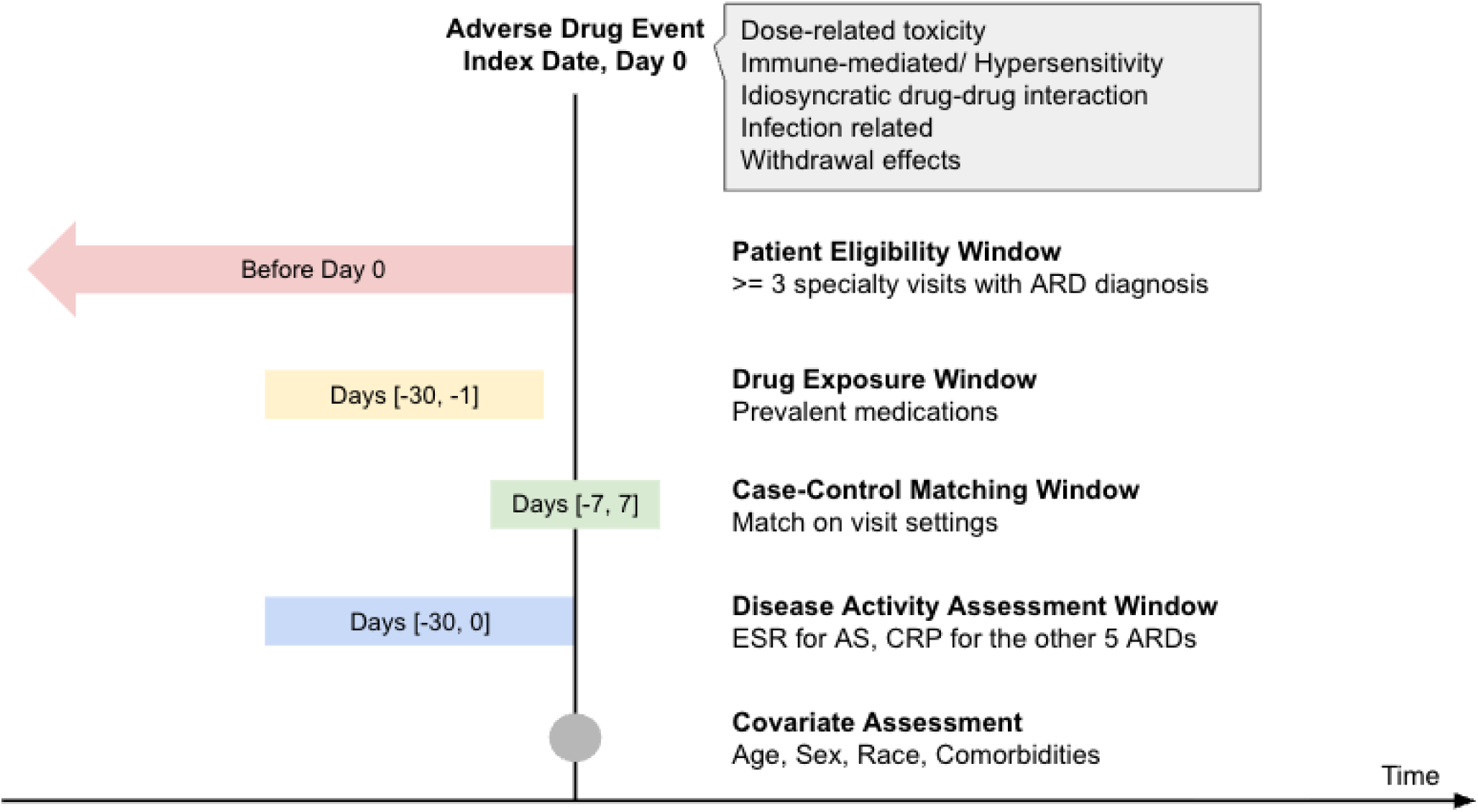
Study Timeline and Analytic Windows for the Nested Encounter-Matched Case–Control Analysis of Adverse Drug Events

### Study Population and Cohort Definition

We identified adults aged ≥18 years with AS, PsA, RA, SjD, SLE, SSc using validated ICD-9 and ICD-10 code–based definitions (Supplementary Table 1). To improve diagnostic specificity, we required at least three outpatient visits with an ARD diagnosis recorded in relevant specialty clinics (rheumatology, nephrology, or dermatology).^13^

### ADE outcome definition

ADEs were defined as medication-related clinical events occurring in outpatient, inpatient, or emergency department encounters after ARD diagnosis. We restricted analyses to the first ADE per patient. ADEs were identified using a validated ICD-10–based algorithm informed by the Hohl framework,^14^ incorporating categories A–C and selected category D codes with strong pharmacologic specificity (Supplementary Table 2). This ICD-based ADE definition prioritizes specificity (85–95%) at the expense of sensitivity (3-28%), consistent with prior EHR-based ADE studies.^14,15^

### Medication exposure definition

Medications were harmonized using the Anatomical Therapeutic Chemical classification system^16^ to ensure consistent grouping across therapeutic classes. Prescription records were used to construct exposure episodes, with overlapping or consecutive prescriptions merged to represent continuous use. A medication was considered active if its exposure window overlapped the 30 days preceding the index date (ADE date for cases; matched encounter date for controls). Polypharmacy was defined as concurrent use of ≥5 medications within a 30-day window and categorized as minor (5–10) or major (>10). Equivalent formulations were deduplicated at the ingredient and class level.

### Covariates

Demographic variables included age, sex, race and ethnicity, body mass index (BMI), smoking status, and insurance type at the index date. Comorbidities were identified using ICD codes within the prior 12 months and summarized using the Charlson Comorbidity Index (CCI).^17^

### Descriptive analysis

We compared patient characteristics, comorbidities, medication use, and healthcare utilization between ADE and non-ADE groups at the index date. Continuous variables were summarized using means with standard deviations (SD) or medians with interquartile ranges (IQR) and compared using t-tests or Wilcoxon rank-sum tests. Categorical variables were summarized as counts and percentages and compared using chi-square or Fisher’s exact tests.

### Case-control analysis

Within each ARD cohort, we conducted a nested case–control analysis to evaluate associations between medication exposure and ADEs. Cases were encounters with ADEs, and up to four control encounters without ADEs were matched on ARD diagnosis and encounter type (outpatient, inpatient, or emergency department). Matching at the encounter level allowed individuals to contribute multiple control encounters while preserving temporal alignment and minimizing immortal time bias.

For each case and control, we identified active medications in the 30 days preceding the index date. We evaluated polypharmacy burden and examined exposure to commonly used medications. Associations were estimated using conditional logistic regression, adjusting for age, sex, race and ethnicity, and CCI. Results are reported as adjusted odds ratios (aOR) with 95% confidence intervals, stratified by ARD.

### ADE code prevalence

To characterize ADE patterns, we examined the distribution of ICD-10 codes among ADE cases. To reduce sparsity, we identified the 30 most frequent ADE-related codes in RA (the largest cohort) and evaluated their prevalence across all ARDs. The reported mean proportion of T codes (30.0%), for example, reflects the proportion of T codes among the top 30 ADE codes within each ARD, averaged across all six diseases.

### Sensitivity analysis

To assess potential confounding by disease activity, we incorporated disease-specific biomarkers measured within 30 days before the index date. Biomarker selection was guided by two rheumatologists (CA and MW) and literature review. C-reactive protein (CRP) or erythrocyte sedimentation rate (ESR) was used depending on disease and data availability. Analyses were repeated in subsets with available biomarker data to evaluate robustness of effect estimates.

## RESULTS

### Demographic and clinical correlates

We identified 10,578 patients with ARDs between 2010 and 2024, of whom 3,154 (29.8%) experienced at least one ADE (Table 1). The cohort was predominantly female (76.6%) and White (61.8%), with representation of Asian (20.1%) and Black (2.9%) patients. Polypharmacy was highly prevalent, with 57.3% of patients exposed to 5–10 medications and 39.4% exposed to more than 10. RA comprised the largest subgroup (38.2%), followed by PsA (17.6%), SLE (16.8%), SjD (15.1%), SSc (7.6%), and AS (4.9%).

**Table 1.** Baseline characteristics of the pooled study population across six ARD conditions.

| Characteristic | Overall (N=10,578) | ADE Yes (N=3,154) | ADE No (N=7,424) |
| --- | --- | --- | --- |
| **Sex** |  |  |  |
| Male | 2,474 (23.4%) | 643 (26.0%) | 1,831 (74.0%) |
| Female | 8,104 (76.6%) | 2,511 (31.0%) | 5,593 (69.0%) |
| **Age, years** |  |  |  |
| Mean (SD)* | 56.0 (17.2) | 58.2 (17.1) | 55.1 (17.1) |
| **Race** |  |  |  |
| White | 6,540 (61.8%) | 2,106 (32.2%) | 4,434 (67.8%) |
| Asian | 2,125 (20.1%) | 512 (24.1%) | 1,613 (75.9%) |
| Black | 306 (2.9%) | 116 (37.9%) | 190 (62.1%) |
| Other | 145 (1.4%) | 50 (34.5%) | 95 (65.5%) |
| Unknown | 1,462 (13.8%) | 370 (25.3%) | 1,092 (74.7%) |
| **Ethnicity** |  |  |  |
| Hispanic/Latino | 1,329 (12.6%) | 419 (31.5%) | 910 (68.5%) |
| Non-Hispanic | 8,742 (82.6%) | 2,643 (30.2%) | 6,099 (69.8%) |
| Unknown | 507 (4.8%) | 91 (17.9%) | 416 (82.1%) |
| **Insurance** |  |  |  |
| Private | 5,377 (50.8%) | 1,468 (27.3%) | 3,909 (72.7%) |
| Public | 1,866 (17.6%) | 647 (27.4%) | 1,718 (72.6%) |
| Unknown | 3,335 (31.5%) | 1,039 (31.2%) | 2,296 (68.8%) |
| **Recent BMI** |  |  |  |
| Mean (SD)* | 26.8 (6.4) | 26.9 (6.5) | 26.7 (6.3) |
| Missing | 137 (1.3%) | 5 (0.2%) | 132 (1.8%) |
| **Smoking status** |  |  |  |
| No | 8,238 (77.9%) | 2,261 (27.4%) | 5,977 (72.6%) |
| Yes | 2,321 (21.9%) | 884 (38.1%) | 1,437 (61.9%) |
| Unknown | 19 (0.2%) | 9 (47.4%) | 10 (52.6%) |
| **Charlson Comorbidity Index (0–17), Mean (SD)\*** | 2.28 (2.04) | 3.47 (2.44) | 1.86 (1.58) |
| **Polypharmacy** |  |  |  |
| Minor (5-10 drugs) | 6,059 (57.3%) | 2,290 (37.8%) | 3,769 (62.2%) |
| Major (>10 drugs) | 4,167 (39.4%) | 1,876 (45.0%) | 2,291 (55.0%) |
| **Disease** |  |  |  |
| Ankylosing spondylitis | 514 (4.9%) | 153 (29.7%) | 361 (70.2%) |
| Psoriatic arthritis | 1859 (17.6%) | 532 (28.6%) | 1327 (71.4%) |
| Rheumatoid arthritis | 4036 (38.2%) | 1146 (28.4%) | 2890 (71.6%) |
| Sjögren's disease | 1596 (15.1%) | 499 (31.3%) | 1097 (68.7%) |
| Systemic Lupus Erythematosus | 1774 (16.8%) | 537 (30.3) | 1237 (69.7%) |
| Systemic sclerosis | 799 (7.6%) | 287 (35.9%) | 512 (64.1%) |

Patients with ADEs were slightly older (mean 58.2 vs 55.1 years, Table 1) and had higher comorbidity burden (CCI 3.47 vs 1.86) than those without ADEs. ADEs were more frequent among females (31.0% vs. 26.0%), Black (37.9% vs. White 32.2%) and smokers (38.1% vs 27.4%). Differences by insurance type and body mass index were minimal.

### Medication Burden Across Care Settings

In the matched case–control analysis, ADE cases consistently had greater medication burden than controls across all ARDs (Supplementary Table 3). This pattern was observed in outpatient, inpatient, and emergency department encounters. For example, among patients with SLE, median medication counts were approximately two-fold higher in cases compared with controls across all settings (17 vs. 8 in emergency, 20 vs. 10 in inpatients, 12 vs.6 in outpatient). Similar differences were observed across other ARDs, indicating a robust association between polypharmacy and ADE risk.

### Medication and ADE Associations

Associations between medications and ADEs were heterogeneous across diseases and drug classes (Figure 2; Supplementary Tables 4-9). Overall, the strongest associations were observed for supportive and symptom-directed therapies, whereas disease-modifying treatments generally showed weaker or inverse associations. Gastrointestinal agents, particularly acid-suppressive therapies, demonstrated consistently elevated associations across multiple ARDs. This pattern was most prominent in SSc (pantoprazole aOR 4.12 [2.56-6.65], famotidine aOR 4.92 [3.06-7.92]), with additional strong signals observed for prokinetic agents (metoclopramide aOR 12.32 [6.99-21.71]). Analgesics and pain adjuncts were also consistently associated with ADEs across diseases, with higher effect estimates observed in SLE (hydromorphone aOR 7.97 [5.60-11.34], oxycodone aOR 6.05 [4.31-8.05], acetaminophen aOR 9.68 [5.63-16.66]) and SSc (hydromorphone aOR 5.46 [3.43-8.68], oxycodone aOR 2.97 [1.92-4.58], acetaminophen aOR 7.21 [3.52-14.77]).

**Figure 2.**
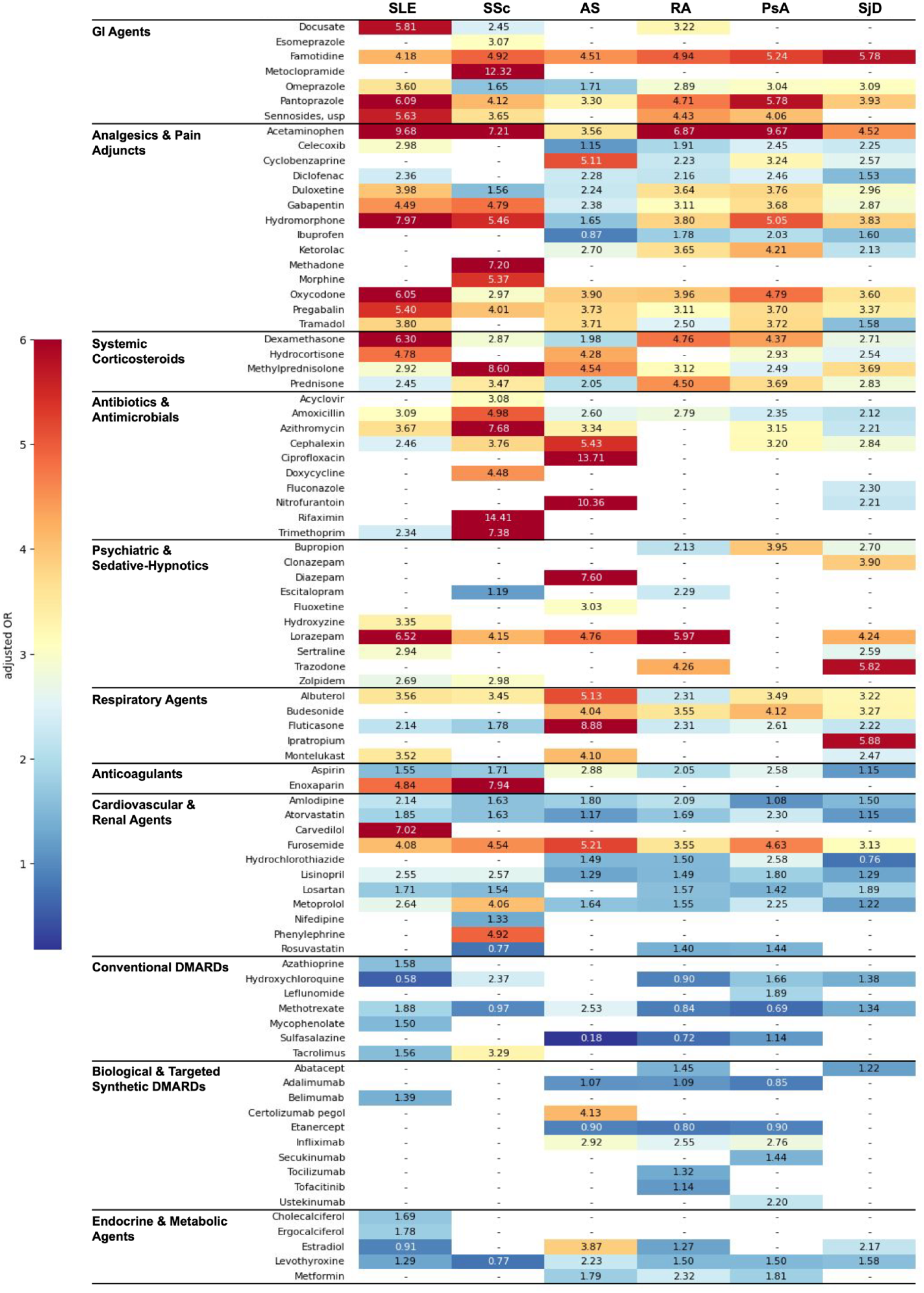
Heatmap of adjusted odds ratios for the top 50 medications associated with adverse drug events, stratified by drug class across six ARD conditions.

Systemic corticosteroids showed strong associations in selected diseases, particularly SLE (dexamethasone aOR 6.30 [4.48-8.86], methylprednisolone aOR 2.92 [2.15-3.96]) and SSc (dexamethasone aOR 2.87 [1.82-4.54], methylprednisolone aOR 8.60 [5.08-14.56]), while associations were weaker or absent in other ARDs. Antibiotics and antimicrobials exhibited disease-specific patterns rather than uniform effects, specifically in SSc (azithromycin aOR 7.68 [4.49-13.14]) and AS (cephalexin aOR 5.43 [2.98-9.91]). Psychiatric and sedative-hypnotic medications demonstrated heterogeneous but generally elevated associations across several ARDs.

Respiratory medications showed prominent associations in AS (fluticasone aOR 8.88 [4.44-17.75], albuterol aOR 5.13 [2.70-9.75]), whereas cardiovascular and renal agents exhibited modest but widespread associations. In contrast, DMARDs and endocrine or metabolic medications were generally associated with lower or inconsistent effect estimates.

### Disease-Specific Patterns

Disease-stratified analyses revealed both consistent and distinct medication patterns across ARDs (Supplementary Tables 4-9; Figure 2). Across six ARDs, analgesics, sedatives, gastrointestinal agents, corticosteroids and antimicrobials were recurrently associated with ADEs.

SLE (Supplementary Table 4) and RA (Supplementary Table 5) showed similar profiles, with strong associations for analgesic and sedative medications and weaker or inverse associations for selected DMARDs. PsA (Supplementary Table 6) demonstrated a broadly similar pattern, with multiple symptomatic medications associated with ADEs and weaker associations for DMARDs. SSc (Supplementary Table 7) demonstrated particularly strong associations with gastrointestinal agents. AS (Supplementary Table 8) was characterized by stronger associations with antibiotics and respiratory medications. SjD (Supplementary Table 9) showed a mixed pattern involving respiratory, gastrointestinal, and psychotropic medications.

### ADE Code Distribution

The diversity of ADE manifestations varied across ARDs, with the number of unique ICD-10 codes ranging from 45 in AS to 152 in RA. Using the 30 most frequent ADE codes in RA as a reference set (Supplementary Table 10), substantial overlap was observed in the other five diseases (67–87%).

Across ARDs, ADEs were most frequently represented by external cause codes (ICD chapter T, 30.0%) and digestive system codes (ICD chapter K, 25.6%), followed by neurologic (ICD chapter G, 8.9%), psychiatric (ICD chapter F, 8.3%), dermatologic (ICD chapter L, 7.2%), endocrine (ICD chapter E, 6.7%), infectious (ICD chapter A, 4.4%), and cardiovascular categories (ICD chapter I, 3.3%). ICD chapters were defined by the first character of the diagnosis code (e.g., T codes for external causes, K codes for digestive system conditions). This pattern suggests that gastrointestinal manifestations are the most common ADE presentations.

### Sensitivity Analyses

Adjustment for demographic and comorbidity variables increased effect estimates relative to unadjusted models. Further adjustment for disease activity generally attenuated associations (Supplementary Table 4-9), suggesting partial confounding by disease activity. After adjustment for demographics, comorbidity burden, and disease activity, several medication associations strengthened modestly. For example, in SLE, aORs increased for tacrolimus (1.56 to 1.90) and belimumab (1.39 to 2.06), while in RA, tocilizumab shifted from a non-significant association (1.32 [0.93-1.90]) to a significant association (1.70 [1.31-2.21]). Similar patterns were observed across other ARDs, including prednisone in SSc (3.47 to 4.22), topiramate in AS (2.09 to 4.70), and valacyclovir in SjD (1.70 to 2.30).

## DISCUSSION

In this large EHR-based study of 10,578 patients across six ARDs, nearly one-third experienced an ADE. ADE burden varied across diseases, with the highest prevalence observed in SSc (35.9%). Polypharmacy was highly prevalent across all diseases and care settings, and ADE risk consistently increased with greater concurrent medication exposure. Notably, ADE signals were concentrated in symptom-directed therapies rather than DMARDs. These patterns were shared across ARDs but also demonstrated disease-specific variation, including gastrointestinal agents in SSc and respiratory and antibiotic therapies in AS.

Our findings reinforce that polypharmacy is a structural feature of ARD care. Prior studies in RA and SLE have reported polypharmacy prevalence exceeding 60–70%, substantially higher than in the general population^9,10^. We extend this literature by demonstrating that polypharmacy is not only prevalent but also consistently associated with ADE risk across multiple ARDs and clinical settings within a large real-world cohort. This relationship was graded, with ADE cases exhibiting approximately two-fold higher medication counts than matched controls in several settings, consistent with a dose–response association between medication burden and ADE risk.^18,19^ Importantly, this association was observed within matched case-control comparisons, suggesting that differences in medication burden are not solely attributable to underlying disease distribution.

A central finding is that ADE risk was more strongly associated with supportive and symptom-directed medications, including gastrointestinal agents, analgesics, and psychotropic therapies, than with DMARDs. Although these associations are not causal, this pattern is biologically and clinically coherent. Effective control of inflammatory disease activity can reduce reliance on symptomatic medications, whereas persistent symptoms often lead to layered prescribing and cumulative exposure. In contrast, DMARD exposure may reflect more stable disease management and closer clinical monitoring,^20^ with reduced need for symptomatic medications and lower observed ADE risk. However, this likely reflects confounding by disease activity; adjustment for disease activity attenuated these associations, although they remained present.

Analgesics and pain adjuncts emerged as a particularly strong and clinically actionable signal. Pain remains prevalent in ARDs even in the setting of controlled inflammation,^21^ and management frequently involves sequential or concurrent use of multiple agents.^22^ This layered prescribing increases the risk of adverse outcomes through additive central nervous system depression, renal and gastrointestinal toxicity, and increased fall risk.^23^ The consistent association of analgesic and sedative medications with ADEs across diseases suggests that these therapies may reflect higher disease activity and cumulative medication exposure, as well as contribute to risk. Taking acetaminophen as an example, despite its favorable safety profile, its use likely reflects greater symptom burden and treatment intensity rather than direct toxicity, indicating that patients with poorly controlled disease accumulate symptomatic therapies and higher ADE risk. Systemic corticosteroids also demonstrated strong associations with ADEs, particularly in SLE and SSc. This finding is consistent with established evidence linking corticosteroid exposure to infection, metabolic complications, and hospitalization.^12,24^ The magnitude and consistency of these associations reinforce the importance of steroid minimization strategies and proactive risk mitigation in routine care. Despite shared patterns, we observed disease-specific differences in associations between medications and ADEs. In SSc, ADE-associated medications were concentrated in gastrointestinal therapies, consistent with the high prevalence of gastrointestinal involvement and dysmotility.^25^ In AS, respiratory medications and antibiotics were more prominent, aligning with increased susceptibility to infection-related morbidity.^26^ These disease-specific signatures suggest that ADE risk is shaped not only by medication burden but also by organ-specific disease manifestations and treatment patterns.

SSc appears under-recognized yet disproportionately impacted, with the highest ADE burden observed in our analysis. SSc is a complex multisystem disease characterized by substantial morbidity driven by interstitial lung disease, pulmonary hypertension, and gastrointestinal involvement, often requiring intensive and overlapping pharmacotherapy.^27^ Prior studies similarly report high medication burden and drug–drug interaction risk in SSc populations.^8^ Pharmacokinetic vulnerability may further contribute; for example, co-administration of acid-suppressive therapies has been shown to reduce mycophenolate mofetil bioavailability.^28^ Our findings suggest SSc represents a high-risk phenotype for ADEs and may benefit from targeted surveillance and deliberate medication optimization.

Across ICD-based ADE classifications, gastrointestinal manifestations were among the most common clinical presentations, second only to external cause codes. This pattern is consistent with the pharmacologic profiles of commonly used medications in ARDs, including NSAIDs and corticosteroids,^29^ and suggests that the gastrointestinal system represents a major site of ADEs in this population.

A key methodological challenge in our sensitivity analyses was the selection of appropriate biomarkers to represent disease activity across ARDs. Because no single marker uniformly captures inflammatory burden in all diseases, we balanced clinical expertise, strength of published evidence, and data availability in our cohort. Incorporating these biomarkers led to overall attenuation of associations, suggesting that part of the crude effects was attributable to confounding by inflammatory activity, while selective strengthening of certain medication associations indicates heterogeneity in confounding structure across drugs and disease phenotypes. However, biomarker data were available for only 30–47% of patients in each ARD cohort, reducing sample size and potentially limiting statistical power, and single measurements within 30 days may not fully capture longitudinal disease activity. These constraints underscore the challenges of adequately adjusting for disease activity in large EHR-based studies.

Several additional limitations should be considered. First, ADE identification relied on ICD-based algorithms, which prioritize specificity but may undercapture less severe or undocumented events.^30^ Second, medication exposure was defined using a 30-day pre-index window, which may not fully capture the biologically relevant risk period for all drugs, particularly those with long-term effects or prolonged pharmacodynamic activity (e.g., rituximab), and may introduce exposure misclassification. Third, we did not explicitly model drug–drug interactions, a key mechanism underlying polypharmacy-related harm, potentially underestimating synergistic risks. Fourth, residual confounding remains possible. We did not directly adjust for healthcare utilization intensity, and ARDs disease severity was incompletely captured within a single-center EHR; although we used the CCI as a proxy, it may not fully reflect underlying disease burden or care complexity. Finally, important contextual determinants of prescribing, such as socioeconomic factors, care fragmentation, prescriber type, and access to medications, were not captured, limiting insight into upstream drivers of polypharmacy.

Building on the consistent cross-disease patterns observed in this large, real-world cohort, several next steps could enhance clinical translation. First, the robust association between medication burden, particularly supportive and symptom-directed therapies, and ADE risk provides a foundation for developing targeted risk stratification tools embedded within the EHRs, enabling identification of high-risk patients for proactive medication review. Second, the disease-specific medication signatures identified across ARDs suggest an opportunity to move beyond generic polypharmacy metrics toward more tailored, disease-informed medication optimization strategies. Embedding these insights into structured medication review processes, such as identifying high-risk combinations during routine visits, may support targeted deprescribing and safer prescribing decisions. Third, prospective studies across diverse healthcare settings are needed to determine whether modifying high-risk prescribing patterns, particularly in pain management and corticosteroid use, reduces ADE burden and to evaluate the generalizability and implementation of these strategies in routine care.

In summary, ADE risk in ARDs reflects both underlying disease activity and cumulative exposure to symptom-directed medications, with both shared and disease-specific patterns across diseases. While disease activity contributes to observed associations, our findings suggest that medication burden, particularly from supportive therapies, represents a key and potentially modifiable contributor to harm. Strategies focused on optimizing prescribing, minimizing corticosteroid exposure, and improving pain management can reduce ADE burden in this population.

## Data Availability

Aggregate data produced in the present study are contained in the manuscript and supplementary materials. Individual-level electronic health record data are not publicly available because of privacy, ethics, and institutional data-use restrictions.

---

**Supplementary Table 1.** Case Definition for Autoimmune Rheumatic Diseases (ARDs)

| Condition | ICD-9 Codes | ICD-10 Codes | Specialist |
| --- | --- | --- | --- |
| Ankylosing spondylitis | 720.0 | M45.X | Rheumatologist |
| Psoriatic arthritis | 696.0 | L40.5X | Rheumatologist,<br>Dermatologist |
| Rheumatoid arthritis | 714.0, 714.1, 714.2,<br>714.8 | M05.X, M06.X | Rheumatologist |
| Sjögren's disease | 710.2 | M35.0X | Rheumatologist |
| Systemic lupus erythematosus | 710.0 | M32.X (excluding<br>M32.0) | Rheumatologist,<br>Dermatologist,<br>Nephrologist |
| Systemic sclerosis | 710.1 | M34.X | Rheumatologist,<br>Dermatologist |

**Supplementary Table 2.** ICD-10 codes identifying adverse drug events classified by Hohl causality category.

| Hohl Category | ICD-10 Code | Description |
| --- | --- | --- |
| A1 — Induced by medication | D59.0 | Drug-induced autoimmune haemolytic anaemia |
| A1 — Induced by medication | D61.1 | Drug-induced aplastic anaemia due to chemotherapy |
| A1 — Induced by medication | D68.312 | Hemorrhagic disorder due to circulating anticoagulants |
| A1 — Induced by medication | D68.318 | Hemorrhagic disorder due to circulating anticoagulants |
| A1 — Induced by medication | D68.32 | Hemorrhagic disorder due to circulating anticoagulants |
| A1 — Induced by medication | E16.0,<br>T38.3X5A | Drug-induced hypoglycaemia without coma |
| A1 — Induced by medication | E24.2 | Drug-induced Cushing's syndrome |
| A1 — Induced by medication | E27.3 | Drug-induced adrenocortical insufficiency |
| A1 — Induced by medication | E27.3,<br>T38.0X5A | Drug-induced adrenocortical insufficiency |
| A1 — Induced by medication | E66.1 | Drug-induced obesity |
| A1 — Induced by medication | G25.1 | Drug-induced tremor |
| A1 — Induced by medication | G44.40 | Drug-induced headache, not elsewhere classified |
| A1 — Induced by medication | G44.40,<br>T39.95XA | Drug-induced headache, not elsewhere classified |
| A1 — Induced by medication | G44.41 | Drug-induced headache, not elsewhere classified |
| A1 — Induced by medication | G62.0 | Drug-induced polyneuropathy |
| A1 — Induced by medication | G72.0 | Drug-induced myopathy |
| A1 — Induced by medication | G72.0,<br>T38.0X5A | Drug-induced myopathy |
| A1 — Induced by medication | G72.0,<br>T46.6X5A | Drug-induced myopathy |
| A1 — Induced by medication | H26.31 | Drug-induced cataract |
| A1 — Induced by medication | H26.33 | Drug-induced cataract |
| A1 — Induced by medication | H40.60X0 | Glaucoma secondary to drugs |
| A1 — Induced by medication | H40.63X0 | Glaucoma secondary to drugs |
| A1 — Induced by medication | H40.63X0,<br>T38.0X5A | Glaucoma secondary to drugs |
| A1 — Induced by medication | I95.2 | Hypotension due to drugs |
| A1 — Induced by medication | K85.30 | Drug-induced pancreatitis |
| A1 — Induced by medication | L43.2 | Lichenoid drug reaction |
| A1 — Induced by medication | M87.10 | Osteonecrosis due to drugs |
| A1 — Induced by medication | M87.10,<br>T38.0X5A | Osteonecrosis due to drugs |
| A1 — Induced by medication | M87.111,<br>T38.0X5A | Osteonecrosis due to drugs |
| A1 — Induced by medication | M87.119,<br>T38.0X5A | Osteonecrosis due to drugs |
| A1 — Induced by medication | M87.150 | Osteonecrosis due to drugs |
| A1 — Induced by medication | M87.151 | Osteonecrosis due to drugs |
| A1 — Induced by medication | M87.152 | Osteonecrosis due to drugs |
| A1 — Induced by medication | M87.161 | Osteonecrosis due to drugs |
| A1 — Induced by medication | M87.162 | Osteonecrosis due to drugs |
| A1 — Induced by medication | M87.174 | Osteonecrosis due to drugs |
| A1 — Induced by medication | M87.177 | Osteonecrosis due to drugs |
| A1 — Induced by medication | M87.180 | Osteonecrosis due to drugs |
| A1 — Induced by medication | M87.180,<br>T45.8X5A | Osteonecrosis due to drugs |
| A1 — Induced by medication | M87.19 | Osteonecrosis due to drugs |
| A1 — Induced by medication | P04.18, Z91.B | Fetus and newborn affected by other maternal medication |
| A1 — Induced by medication | R50.2 | Drug-induced fever |
| A2 — Induced by medication or other causes | E03.2 | Hypothyroidism due to medicaments and other exogenous substances |
| A2 — Induced by medication or other causes | F11.10 | Mental and behavioural disorders due to use of opioids:<br>Harmful use |
| A2 — Induced by medication or other causes | F11.20 | Mental and behavioural disorders due to use of opioids:<br>Dependence syndrome |
| A2 — Induced by medication or other causes | F11.23 | Mental and behavioural disorders due to use of opioids:<br>Dependence syndrome |
| A2 — Induced by medication or other causes | F11.288 | Mental and behavioural disorders due to use of opioids:<br>Dependence syndrome |
| A2 — Induced by medication or other causes | F11.29 | Mental and behavioural disorders due to use of opioids:<br>Dependence syndrome |
| A2 — Induced by medication or other causes | F11.90 | Mental and behavioural disorders due to use of opioids:<br>Unspecified mental and behavioural disorder |
| A2 — Induced by medication or other causes | F11.93 | Mental and behavioural disorders due to use of opioids:<br>Unspecified mental and behavioural disorder |
| A2 — Induced by medication or other causes | F11.99 | Mental and behavioural disorders due to use of opioids: Unspecified mental and behavioural disorder |
| A2 — Induced by medication or other causes | F13.10 | Mental and behavioural disorders due to use of sedatives or hypnotics: Harmful use |
| A2 — Induced by medication or other causes | F13.20 | Mental and behavioural disorders due to use of sedatives or hypnotics: Dependence syndrome |
| A2 — Induced by medication or other causes | F15.10 | Mental and behavioural disorders due to use of other stimulants, including caffeine: Harmful use |
| A2 — Induced by medication or other causes | F15.21 | Mental and behavioural disorders due to use of other stimulants, including caffeine: Dependence syndrome |
| A2 — Induced by medication or other causes | F19.10 | Mental and behavioural disorders due to multiple drug use and use of other psychoactive substances: Harmful use |
| A2 — Induced by medication or other causes | F19.11 | Mental and behavioural disorders due to multiple drug use and use of other psychoactive substances: Harmful use |
| A2 — Induced by medication or other causes | F19.20 | Mental and behavioural disorders due to multiple drug use and use of other psychoactive substances: Dependence syndrome |
| A2 — Induced by medication or other causes | F19.21 | Mental and behavioural disorders due to multiple drug use and use of other psychoactive substances: Dependence syndrome |
| A2 — Induced by medication or other causes | F19.239 | Mental and behavioural disorders due to multiple drug use and use of other psychoactive substances: Dependence syndrome |
| A2 — Induced by medication or other causes | F19.90 | Mental and behavioural disorders due to multiple drug use and use of other psychoactive substances: Unspecified mental and behavioural disorder |
| A2 — Induced by medication or other causes | F19.939 | Mental and behavioural disorders due to multiple drug use and use of other psychoactive substances: Unspecified mental and behavioural disorder |
| A2 — Induced by medication or other causes | F19.951 | Mental and behavioural disorders due to multiple drug use and use of other psychoactive substances: Unspecified mental and behavioural disorder |
| A2 — Induced by medication or other causes | F19.982 | Mental and behavioural disorders due to multiple drug use and use of other psychoactive substances: Unspecified mental and behavioural disorder |
| A2 — Induced by medication or other causes | L25.1 | Unspecified contact dermatitis due to drugs in contact with skin |
| A2 — Induced by medication or other causes | L27.0 | Generalized skin eruption due to drugs and medicaments |
| A2 — Induced by medication or other causes | L27.0, T36.1X5A | Generalized skin eruption due to drugs and medicaments |
| A2 — Induced by medication or other causes | L27.1 | Localized skin eruption due to drugs and medicaments |
| A2 — Induced by medication or other causes | L27.2 | Dermatitis due to substances taken internally |
| A2 — Induced by medication or other causes | N14.1 | Nephropathy induced by other drugs, medicaments and biological substances |
| A2 — Induced by medication or other causes | N14.11 | Nephropathy induced by other drugs, medicaments and biological substances |
| A2 — Induced by medication or other causes | N14.11, T50.8X5A | Nephropathy induced by other drugs, medicaments and biological substances |
| A2 — Induced by medication or other causes | N14.19 | Nephropathy induced by other drugs, medicaments and biological substances |
| A2 — Induced by medication or other causes | N14.19, T45.1X5A | Nephropathy induced by other drugs, medicaments and biological substances |
| A2 — Induced by medication or other causes | N14.4 | Toxic nephropathy, not elsewhere classified |
| A2 — Induced by medication or other causes | T78.00XA | Adverse effects, not elsewhere classified |
| A2 — Induced by medication or other causes | T78.00XD | Adverse effects, not elsewhere classified |
| A2 — Induced by medication or other causes | T78.00XS | Adverse effects, not elsewhere classified |
| A2 — Induced by medication or other causes | T78.05XA | Adverse effects, not elsewhere classified |
| A2 — Induced by medication or other causes | T78.19XA | Adverse effects, not elsewhere classified |
| A2 — Induced by medication or other causes | T78.19XD | Adverse effects, not elsewhere classified |
| A2 — Induced by medication or other causes | T78.1XXA | Adverse effects, not elsewhere classified |
| A2 — Induced by medication or other causes | T78.1XXD | Adverse effects, not elsewhere classified |
| A2 — Induced by medication or other causes | T78.2XXA | Anaphylactic shock, unspecified |
| A2 — Induced by medication or other causes | T78.2XXD | Anaphylactic shock, unspecified |
| A2 — Induced by medication or other causes | T78.2XXS | Anaphylactic shock, unspecified |
| A2 — Induced by medication or other causes | T78.3XXA | Angioneurotic oedema |
| A2 — Induced by medication or other causes | T78.3XXD | Angioneurotic oedema |
| A2 — Induced by medication or other causes | T78.3XXS | Angioneurotic oedema |
| A2 — Induced by medication or other causes | T78.40XA | Allergy, unspecified |
| A2 — Induced by medication or other causes | T78.40XA,<br>R06.2 | Allergy, unspecified |
| A2 — Induced by medication or other causes | T78.40XA,<br>R21 | Allergy, unspecified |
| A2 — Induced by medication or other causes | T78.40XD | Allergy, unspecified |
| A2 — Induced by medication or other causes | T78.40XS | Allergy, unspecified |
| A2 — Induced by medication or other causes | T78.49XA | Allergy, unspecified |
| A2 — Induced by medication or other causes | T78.49XD,<br>T39.8X5D | Allergy, unspecified |
| A2 — Induced by medication or other causes | T80.211A | Infections following infusion, transfusion and therapeutic injection |
| A2 — Induced by medication or other causes | T80.211D | Infections following infusion, transfusion and therapeutic injection |
| A2 — Induced by medication or other causes | T80.212A | Infections following infusion, transfusion and therapeutic injection |
| A2 — Induced by medication or other causes | T80.218A | Infections following infusion, transfusion and therapeutic injection |
| A2 — Induced by medication or other causes | T80.219A | Infections following infusion, transfusion and therapeutic injection |
| A2 — Induced by medication or other causes | T80.52XA | Complications following infusion, transfusion and therapeutic injection: anaphylactic shock due to serum |
| A2 — Induced by medication or other causes | T80.62XA | Complications following infusion, transfusion and therapeutic injection: other serum reactions |
| A2 — Induced by medication or other causes | T80.69XA | Complications following infusion, transfusion and therapeutic injection: other serum reactions |
| A2 — Induced by medication or other causes | T80.810A | Other complications following infusion, transfusion and therapeutic injection |
| A2 — Induced by medication or other causes | T80.818A | Other complications following infusion, transfusion and therapeutic injection |
| A2 — Induced by medication or other causes | T80.818A, I96 | Other complications following infusion, transfusion and therapeutic injection |
| A2 — Induced by medication or other causes | T80.82XA | Other complications following infusion, transfusion and therapeutic injection |
| A2 — Induced by medication or other causes | T80.89XA | Other complications following infusion, transfusion and therapeutic injection |
| A2 — Induced by medication or other causes | T80.89XA,<br>T88.7XXA | Other complications following infusion, transfusion and therapeutic injection |
| A2 — Induced by medication or other causes | T80.90XA | Unspecified complication following infusion, transfusion and therapeutic injection |
| A2 — Induced by medication or other causes | T80.90XD | Unspecified complication following infusion, transfusion and therapeutic injection |
| A2 — Induced by medication or other causes | T80.90XS | Unspecified complication following infusion, transfusion and therapeutic injection |
| A2 — Induced by medication or other causes | T80.92XA | Unspecified complication following infusion, transfusion and therapeutic injection |
| A2 — Induced by medication or other causes | T88.6XXA | Anaphylactic shock due to adverse effect of correct drug or medicament properly administered |
| A2 — Induced by medication or other causes | T88.7XXA | Unspecified adverse event of drug or medicament |
| A2 — Induced by medication or other causes | T88.7XXD | Unspecified adverse event of drug or medicament |
| B1 — Poisoning by medication (therapeutic use) | T36.0X5A | Penicillins |
| B1 — Poisoning by medication (therapeutic use) | T36.0X5D | Penicillins |
| B1 — Poisoning by medication (therapeutic use) | T36.1X5A | Cefalosporins and other beta-lactam antibiotics |
| B1 — Poisoning by medication (therapeutic use) | T36.3X5A | Macrolides |
| B1 — Poisoning by medication (therapeutic use) | T36.4X5A | Tetracyclines |
| B1 — Poisoning by medication (therapeutic use) | T36.6X5A | Rifamycins |
| B1 — Poisoning by medication (therapeutic use) | T36.8X5A | Other systemic antibiotics |
| B1 — Poisoning by medication (therapeutic use) | T36.95XA | Systemic antibiotic, unspecified |
| B1 — Poisoning by medication (therapeutic use) | T37.0X5A | Sulfonamides |
| B1 — Poisoning by medication (therapeutic use) | T37.0X5D | Sulfonamides |
| B1 — Poisoning by medication (therapeutic use) | T37.1X5A | Antimycobacterial drugs |
| B1 — Poisoning by medication (therapeutic use) | T37.2X5A | Antimalarials and drugs acting on other blood protozoa |
| B1 — Poisoning by medication (therapeutic use) | T37.5X5A | Antiviral drugs |
| B1 — Poisoning by medication (therapeutic use) | T37.8X1A | Other specified systemic anti-infectives and antiparasitics |
| B1 — Poisoning by medication (therapeutic use) | T37.8X2A | Other specified systemic anti-infectives and antiparasitics |
| B1 — Poisoning by medication (therapeutic use) | T37.8X5A | Other specified systemic anti-infectives and antiparasitics |
| B1 — Poisoning by medication (therapeutic use) | T37.8X5D | Other specified systemic anti-infectives and antiparasitics |
| B1 — Poisoning by medication (therapeutic use) | T37.8X5S | Other specified systemic anti-infectives and antiparasitics |
| B1 — Poisoning by medication (therapeutic use) | T37.8X6A | Other specified systemic anti-infectives and antiparasitics |
| B1 — Poisoning by medication (therapeutic use) | T37.95XA | Systemic anti-infective and antiparasitic, unspecified |
| B1 — Poisoning by medication (therapeutic use) | T38.0X1A | Glucocorticoids and synthetic analogues |
| B1 — Poisoning by medication (therapeutic use) | T38.0X5A | Glucocorticoids and synthetic analogues |
| B1 — Poisoning by medication (therapeutic use) | T38.0X5A,<br>H40.60X0 | Glucocorticoids and synthetic analogues |
| B1 — Poisoning by medication (therapeutic use) | T38.0X5D | Glucocorticoids and synthetic analogues |
| B1 — Poisoning by medication (therapeutic use) | T38.0X5S | Glucocorticoids and synthetic analogues |
| B1 — Poisoning by medication (therapeutic use) | T38.0X6A | Glucocorticoids and synthetic analogues |
| B1 — Poisoning by medication (therapeutic use) | T38.3X5A | Insulin and oral hypoglycaemic [antidiabetic] drugs |
| B1 — Poisoning by medication (therapeutic use) | T38.5X5A | Other estrogens and progestogens |
| B1 — Poisoning by medication (therapeutic use) | T38.895A | Other and unspecified hormones and their synthetic substitutes |
| B1 — Poisoning by medication (therapeutic use) | T39.015A | Salicylates |
| B1 — Poisoning by medication (therapeutic use) | T39.015D | Salicylates |
| B1 — Poisoning by medication (therapeutic use) | T39.1X1A | 4-Aminophenol derivatives |
| B1 — Poisoning by medication (therapeutic use) | T39.1X2A | 4-Aminophenol derivatives |
| B1 — Poisoning by medication (therapeutic use) | T39.1X4A | 4-Aminophenol derivatives |
| B1 — Poisoning by medication (therapeutic use) | T39.395A | Other nonsteroidal anti-inflammatory drugs [NSAID] |
| B1 — Poisoning by medication (therapeutic use) | T39.8X5A | Other nonopioid analgesics and antipyretics, not elsewhere classified |
| B1 — Poisoning by medication (therapeutic use) | T39.95XA | Nonopioid analgesic, antipyretic and antirheumatic, unspecified |
| B1 — Poisoning by medication (therapeutic use) | T40.2X1A | Poisoning by other opioids (includes morphine and codeine) |
| B1 — Poisoning by medication (therapeutic use) | T40.2X5A | Poisoning by other opioids (includes morphine and codeine) |
| B1 — Poisoning by medication (therapeutic use) | T40.605A | Poisoning by other and unspecified narcotics |
| B1 — Poisoning by medication (therapeutic use) | T41.45XA | Anaesthetic, unspecified |
| B1 — Poisoning by medication (therapeutic use) | T42.4X1A | Poisoning by benzodiazepines |
| B1 — Poisoning by medication (therapeutic use) | T42.75XA | Antiepileptic and sedative-hypnotic drugs, unspecified |
| B1 — Poisoning by medication (therapeutic use) | T43.015D | Poisoning by tricyclic and tetracyclic antidepressants |
| B1 — Poisoning by medication (therapeutic use) | T43.205A | Poisoning by other and unspecified antidepressants |
| B1 — Poisoning by medication (therapeutic use) | T43.8X5A | Other psychotropic drugs, not elsewhere classified |
| B1 — Poisoning by medication (therapeutic use) | T44.5X5D | Predominantly beta-adrenoreceptor agonists, not elsewhere classified |
| B1 — Poisoning by medication (therapeutic use) | T44.7X5A | Beta-adrenoreceptor antagonists, not elsewhere classified |
| B1 — Poisoning by medication (therapeutic use) | T45.1X1A | Antineoplastic and immunosuppressive drugs |
| B1 — Poisoning by medication (therapeutic use) | T45.1X1S | Antineoplastic and immunosuppressive drugs |
| B1 — Poisoning by medication (therapeutic use) | T45.1X4A | Antineoplastic and immunosuppressive drugs |
| B1 — Poisoning by medication (therapeutic use) | T45.1X5A | Antineoplastic and immunosuppressive drugs |
| B1 — Poisoning by medication (therapeutic use) | T45.1X5D | Antineoplastic and immunosuppressive drugs |
| B1 — Poisoning by medication (therapeutic use) | T45.1X5D,<br>R11.0 | Antineoplastic and immunosuppressive drugs |
| B1 — Poisoning by medication (therapeutic use) | T45.1X5S | Antineoplastic and immunosuppressive drugs |
| B1 — Poisoning by medication (therapeutic use) | T45.1X6A | Antineoplastic and immunosuppressive drugs |
| B1 — Poisoning by medication (therapeutic use) | T45.4X5A | Iron and its compounds |
| B1 — Poisoning by medication (therapeutic use) | T45.515A | Anticoagulants |
| B1 — Poisoning by medication (therapeutic use) | T45.525A | Anticoagulants |
| B1 — Poisoning by medication (therapeutic use) | T45.8X5A | Other primarily systemic and haematological agents |
| B1 — Poisoning by medication (therapeutic use) | T45.8X6A | Other primarily systemic and haematological agents |
| B1 — Poisoning by medication (therapeutic use) | T46.1X5A | Calcium-channel blockers |
| B1 — Poisoning by medication (therapeutic use) | T46.2X5A | Other antidysrhythmic drugs, not elsewhere classified |
| B1 — Poisoning by medication (therapeutic use) | T46.3X5A | Coronary vasodilators, not elsewhere classified |
| B1 — Poisoning by medication (therapeutic use) | T46.4X5A | Angiotensin-converting-enzyme inhibitors |
| B1 — Poisoning by medication (therapeutic use) | T46.5X5A | Other antihypertensive drugs, not elsewhere classified |
| B1 — Poisoning by medication (therapeutic use) | T46.6X1A | Antihyperlipidaemic and antiarteriosclerotic drugs |
| B1 — Poisoning by medication (therapeutic use) | T46.6X5A | Antihyperlipidaemic and antiarteriosclerotic drugs |
| B1 — Poisoning by medication (therapeutic use) | T46.905A | Other and unspecified agents primarily affecting the cardiovascular system |
| B1 — Poisoning by medication (therapeutic use) | T46.991A | Other and unspecified agents primarily affecting the cardiovascular system |
| B1 — Poisoning by medication (therapeutic use) | T48.6X5A | Antiasthmatics, not elsewhere classified |
| B1 — Poisoning by medication (therapeutic use) | T49.0X1D | Local antifungal, anti-infective and anti-inflammatory drugs, not elsewhere classified |
| B1 — Poisoning by medication (therapeutic use) | T50.2X5A | Carbonic-anhydrase inhibitors, benzothiadiazides and other diuretics |
| B1 — Poisoning by medication (therapeutic use) | T50.4X5A | Drugs affecting uric acid metabolism |
| B1 — Poisoning by medication (therapeutic use) | T50.A95A | Poisoning by diuretics and other unspecified drugs, medicaments and biological substances |
| B1 — Poisoning by medication (therapeutic use) | T50.B95A | Poisoning by diuretics and other unspecified drugs, medicaments and biological substances |
| B1 — Poisoning by medication (therapeutic use) | T50.Z15A | Poisoning by diuretics and other unspecified drugs, medicaments and biological substances |
| B1 — Poisoning by medication (therapeutic use) | T50.Z15D | Poisoning by diuretics and other unspecified drugs, medicaments and biological substances |
| B1 — Poisoning by medication (therapeutic use) | T50.Z95A | Poisoning by diuretics and other unspecified drugs, medicaments and biological substances |
| B1 — Poisoning by medication (therapeutic use) | T50.Z95D | Poisoning by diuretics and other unspecified drugs, medicaments and biological substances |
| B1 — Poisoning by medication (therapeutic use) | T50.Z95S | Poisoning by diuretics and other unspecified drugs, medicaments and biological substances |
| B2 — Poisoning by medication or other causes | T50.8X5A | Diagnostic agents |
| B2 — Poisoning by medication or other causes | T50.8X5D | Diagnostic agents |
| B2 — Poisoning by medication or other causes | T50.901A | Poisoning: Other and unspecified drugs, medicaments and biological substances |
| B2 — Poisoning by medication or other causes | T50.902A | Poisoning: Other and unspecified drugs, medicaments and biological substances |
| B2 — Poisoning by medication or other causes | T50.904A | Poisoning: Other and unspecified drugs, medicaments and biological substances |
| B2 — Poisoning by medication or other causes | T50.905A | Poisoning: Other and unspecified drugs, medicaments and biological substances |
| B2 — Poisoning by medication or other causes | T50.905D | Poisoning: Other and unspecified drugs, medicaments and biological substances |
| B2 — Poisoning by medication or other causes | T50.905S | Poisoning: Other and unspecified drugs, medicaments and biological substances |
| B2 — Poisoning by medication or other causes | T50.906A | Poisoning: Other and unspecified drugs, medicaments and biological substances |
| B2 — Poisoning by medication or other causes | T50.994A | Poisoning: Other and unspecified drugs, medicaments and biological substances |
| B2 — Poisoning by medication or other causes | T50.995A | Poisoning: Other and unspecified drugs, medicaments and biological substances |
| B2 — Poisoning by medication or other causes | T50.995S | Poisoning: Other and unspecified drugs, medicaments and biological substances |
| C — Very likely medication-related | A04.7 | Enterocolitis due to Clostridium difficile |
| C — Very likely medication-related | A04.71 | Enterocolitis due to Clostridium difficile |
| C — Very likely medication-related | A04.72 | Enterocolitis due to Clostridium difficile |
| C — Very likely medication-related | K52.1 | Toxic gastroenteritis and colitis |
| C — Very likely medication-related | K52.1,<br>T36.95XA | Toxic gastroenteritis and colitis |
| C — Very likely medication-related | K71.2 | Toxic liver disease with acute hepatitis |
| C — Very likely medication-related | K71.6 | Toxic liver disease with hepatitis, not elsewhere classified |
| C — Very likely medication-related | K71.6,<br>T50.905A | Toxic liver disease with hepatitis, not elsewhere classified |
| C — Very likely medication-related | K71.9 | Toxic liver disease, unspecified |
| C — Very likely medication-related | L51.1 | Bullous erythema multiforme |
| C — Very likely medication-related | L51.2 | Toxic epidermal necrolysis [Lyell] |
| C — Very likely medication-related | L51.9 | Erythema multiforme, unspecified |
| C — Very likely medication-related | Y69 | Unspecified misadventure during surgical and medical care |
| D — Selected codes likely medication-related | K25.0 | Gastric ulcer: Acute with haemorrhage |
| D — Selected codes likely medication-related | K25.4 | Gastric ulcer: Chronic or unspecified with haemorrhage |
| D — Selected codes likely medication-related | K25.5 | Gastric ulcer: Chronic or unspecified with perforation |
| D — Selected codes likely medication-related | K25.7 | Gastric ulcer: Chronic without haemorrhage or perforation |
| D — Selected codes likely medication-related | K25.9 | Gastric ulcer: Unspecified as acute or chronic, without haemorrhage or perforation |
| D — Selected codes likely medication-related | K26.9 | Duodenal ulcer: Unspecified as acute or chronic, without haemorrhage or perforation |
| D — Selected codes likely medication-related | K27.4 | Peptic ulcer, site unspecified: Chronic or unspecified with haemorrhage |
| D — Selected codes likely medication-related | K27.9 | Peptic ulcer, site unspecified: Unspecified as acute or chronic, without haemorrhage or perforation |
| D — Selected codes likely medication-related | K27.9, B96.81 | Peptic ulcer, site unspecified: Unspecified as acute or chronic, without haemorrhage or perforation |
| D — Selected codes likely medication-related | K28.4 | Gastrojejunal ulcer: Chronic or unspecified with haemorrhage |
| D — Selected codes likely medication-related | K28.9 | Gastrojejunal ulcer: Unspecified as acute or chronic, without haemorrhage or perforation |
| D — Selected codes likely medication-related | K28.9, B96.81 | Gastrojejunal ulcer: Unspecified as acute or chronic, without haemorrhage or perforation |
| D — Selected codes likely medication-related | K29.00 | Acute haemorrhagic gastritis |
| D — Selected codes likely medication-related | K29.01 | Acute haemorrhagic gastritis |
| D — Selected codes likely medication-related | K52.9 | Non-infective gastroenteritis and colitis, unspecified |

**Supplementary Table 3.** Number of medications for ADE cases and controls by visit type.

| Visit Type | Cases*, median (SD) | Controls†, median (SD) |
| --- | --- | --- |
| Systemic lupus erythematosus |  |  |
| Outpatient | 12.0 (11.3) | 6.0 (8.7) |
| Inpatient | 20.0 (14.3) | 10.0 (13.8) |
| Emergency | 17.0 (13.2) | 8.0 (7.7) |
| Rheumatoid arthritis |  |  |
| Outpatient | 11.0 (9.2) | 6.0 (7.3) |
| Inpatient | 14.0 (12.0) | 9.0 (8.0) |
| Emergency | 11.5 (8.7) | 9.0 (9.8) |
| Psoriatic arthritis |  |  |
| Outpatient | 10.0 (10.0) | 6.0 (6.9) |
| Inpatient | 11.5 (8.7) | 10.0 (10.0) |
| Emergency | 12.5 (14.2) | 9.0 (8.0) |
| Systemic sclerosis |  |  |
| Outpatient | 12.5 (12.4) | 6.0 (5.9) |
| Inpatient | 16.5 (12.3) | 10.5 (7.7) |
| Emergency | 15.0 (12.5) | 9.0 (7.1) |
| Ankylosing spondylitis |  |  |
| Outpatient | 9.0 (8.8) | 4.0 (5.5) |
| Inpatient | 19.0 (9.6) | 1.0 (NA) |
| Emergency | 15.0 (8.8) | 7.0 (5.4) |
| Sjögren's disease |  |  |
| Outpatient | 11.0 (10.0) | 6.0 (8.4) |
| Hospital | 14.0 (11.0) | 10.0 (10.5) |
| Emergency | 14.5 (8.5) | 10.0 (12.5) |

\* Medication counts calculated within 30 days prior to the index adverse drug event (ADE) date for cases.
† For matched controls, the same ADE index date from their matched case was used as the reference date

**Supplementary Table 4.** Association between medication exposure and ADEs in SLE.

| Medication | Cases<br>(Exposed<br>/<br>Unexposed) | Controls<br>(Exposed /<br>Unexposed) | Unadjusted OR | Adjusted OR<br>(demographics +<br>comorbidities) | Adjusted OR with<br>ESR (40% of<br>entire SLE cohort) |
| --- | --- | --- | --- | --- | --- |
| acetaminophen | 471 / 66 | 737 / 498 | 6.35 (3.10- 13.02 ) | 9.68 (5.63- 16.66 ) | 4.91 (2.74 - 6.40) |
| hydromorphone | 273 / 264 | 221 / 1014 | 5.23 ( 3.30 - 8.30 ) | 7.97 (5.60- 11.34 ) | 2.58 (1.94 - 3.44) |
| carvedilol | 89 / 448 | 52 / 1183 | 4.59 ( 2.50 - 8.42 ) | 7.02 (4.39- 11.22 ) | 4.52 (3.51 - 5.83) |
| lorazepam | 252 / 285 | 235 / 1000 | 4.41 ( 2.87 - 6.77 ) | 6.52 ( 4.68 - 9.09 ) | 2.51 (1.88 - 3.36) |
| dexamethasone | 294 / 243 | 307 / 928 | 4.30 ( 2.77 - 6.69 ) | 6.30 ( 4.48 - 8.86 ) | 3.66 (3.15 - 4.25) |
| pantoprazole | 283 / 254 | 296 / 939 | 4.21 ( 2.71 - 6.55 ) | 6.09 ( 4.33 - 8.57 ) | 2.45 (1.85 - 3.23) |
| oxycodone | 336 / 201 | 354 / 881 | 4.15 ( 2.67 - 6.45 ) | 6.05 ( 4.31 - 8.50 ) | 4.16 (3.58 - 4.84) |
| docusate | 278 / 259 | 281 / 954 | 4.02 ( 2.56 - 6.29 ) | 5.81 ( 4.10 - 8.22 ) | 2.67 (2.01 - 3.54) |
| sennosides, USP | 240 / 297 | 193 / 1042 | 3.91 ( 2.48 - 6.14 ) | 5.63 ( 3.96 - 8.00 ) | 2.71 (2.03 - 3.63) |
| pregabalin | 115 / 422 | 95 / 1140 | 3.76 ( 2.19 - 6.46 ) | 5.40 ( 3.54 - 8.23 ) | 1.99 (1.37 - 2.91) |
| magnesium sulfate | 191 / 346 | 152 / 1083 | 3.57 ( 2.29 - 5.57 ) | 5.11 ( 3.61 - 7.23 ) | 2.81 (2.07 - 3.81) |
| hydrocortisone | 254 / 283 | 254 / 981 | 3.44 ( 2.24 - 5.27 ) | 4.78 ( 3.42 - 6.67 ) | 2.92 (2.21 - 3.87) |
| enoxaparin | 195 / 342 | 160 / 1075 | 3.43 ( 2.24 - 5.25 ) | 4.84 ( 3.47 - 6.75 ) | 3.06 (2.27 - 4.13) |
| gabapentin | 290 / 247 | 351 / 884 | 3.30 ( 2.13 - 5.09 ) | 4.49 ( 3.20 - 6.31 ) | 1.43 (1.08 - 1.89) |
| famotidine | 270 / 267 | 295 / 940 | 3.07 ( 2.06 - 4.58 ) | 4.18 ( 3.06 - 5.72 ) | 2.03 (1.54 - 2.68) |
| furosemide | 206 / 331 | 218 / 1017 | 3.01 ( 1.99 - 4.55 ) | 4.08 ( 2.95 - 5.66 ) | 2.06 (1.54 - 2.76) |
| duloxetine | 113 / 424 | 138 / 1097 | 2.90 ( 1.76 - 4.79 ) | 3.98 ( 2.68 - 5.92 ) | 1.32 (0.91 - 1.91) |
| tramadol | 178 / 359 | 168 / 1067 | 2.83 ( 1.83 - 4.39 ) | 3.80 ( 2.69 - 5.37 ) | 1.99 (1.46 - 2.71) |
| azithromycin | 244 / 293 | 280 / 955 | 2.79 ( 1.84 - 4.23 ) | 3.67 ( 2.65 - 5.09 ) | 1.93 (1.46 - 2.56) |
| omeprazole | 258 / 279 | 340 / 895 | 2.76 ( 1.82 - 4.16 ) | 3.60 ( 2.61 - 4.98 ) | 1.28 (0.97 - 1.69) |
| albuterol | 284 / 253 | 381 / 854 | 2.67 ( 1.79 - 4.00 ) | 3.56 ( 2.59 - 4.89 ) | 1.56 (1.19 - 2.05) |
| montelukast | 103 / 434 | 102 / 1133 | 2.64 ( 1.62 - 4.31 ) | 3.52 ( 2.39 - 5.19 ) | 2.98 (2.07 - 4.29) |
| hydroxyzine | 178 / 359 | 168 / 1067 | 2.53 ( 1.66 - 3.86 ) | 3.35 ( 2.40 - 4.69 ) | 3.59 (2.67 - 4.83) |
| amoxicillin | 261 / 276 | 338 / 897 | 2.42 ( 1.60 - 3.65 ) | 3.09 ( 2.23 - 4.28 ) | 1.54 (1.17 - 2.02) |
| celecoxib | 124 / 413 | 183 / 1052 | 2.33 ( 1.42 - 3.83 ) | 2.98 ( 2.01 - 4.41 ) | 0.88 (0.61 - 1.28) |
| sertraline | 97 / 440 | 108 / 1127 | 2.33 ( 1.28 - 4.24 ) | 2.94 ( 1.83 - 4.74 ) | 1.36 (0.91 - 2.05) |
| methylprednisolone | 300 / 237 | 390 / 845 | 2.30 ( 1.56 - 3.39 ) | 2.92 ( 2.15 - 3.96 ) | 2.47 (1.86 - 3.28) |
| zolpidem | 130 / 407 | 143 / 1092 | 2.16 ( 1.32 - 3.54 ) | 2.69 ( 1.82 - 3.99 ) | 1.57 (1.08 - 2.29) |
| metoprolol | 168 / 369 | 213 / 1022 | 2.14 ( 1.37 - 3.33 ) | 2.64 ( 1.85 - 3.75 ) | 1.25 (0.91 - 1.72) |
| lisinopril | 139 / 398 | 185 / 1050 | 2.05 ( 1.33 - 3.17 ) | 2.55 ( 1.80 - 3.61 ) | 1.52 (1.10 - 2.09) |
| prednisone | 434 / 103 | 815 / 420 | 2.02 ( 1.26 - 3.22 ) | 2.45 ( 1.69 - 3.55 ) | 2.17 (1.83 - 2.58) |
| cephalexin | 236 / 301 | 292 / 943 | 2.01 ( 1.33 - 3.06 ) | 2.46 ( 1.76 - 3.43 ) | 1.41 (1.06 - 1.88) |
| diclofenac | 196 / 341 | 287 / 948 | 1.96 ( 1.28 - 3.02 ) | 2.36 ( 1.68 - 3.32 ) | 0.82 (0.60 - 1.12) |
| triamcinolone | 334 / 203 | 499 / 736 | 1.94 ( 1.31 - 2.89 ) | 2.35 ( 1.71 - 3.23 ) | 1.50 (1.14 - 1.97) |
| trimethoprim | 209 / 328 | 251 / 984 | 1.92 ( 1.29 - 2.87 ) | 2.34 ( 1.70 - 3.22 ) | 1.78 (1.34 - 2.37) |
| fluticasone | 235 / 302 | 337 / 898 | 1.81 ( 1.24 - 2.64 ) | 2.14 ( 1.58 - 2.89 ) | 1.23 (0.93 - 1.64) |
| amlodipine | 187 / 350 | 243 / 992 | 1.81 ( 1.18 - 2.77 ) | 2.14 ( 1.52 - 3.01 ) | 1.69 (1.26 - 2.27) |
| methotrexate | 132 / 405 | 214 / 1021 | 1.64 ( 1.02 - 2.64 ) | 1.88 ( 1.29 - 2.74 ) | 1.02 (0.74 - 1.42) |
| atorvastatin | 135 / 402 | 213 / 1022 | 1.61 ( 1.02 - 2.54 ) | 1.85 ( 1.29 - 2.67 ) | 1.03 (0.73 - 1.45) |
| ergocalciferol | 198 / 339 | 318 / 917 | 1.56 ( 1.05 - 2.32 ) | 1.78 ( 1.30 - 2.44 ) | 1.95 (1.47 - 2.57) |
| losartan | 122 / 415 | 207 / 1028 | 1.52 ( 0.98 - 2.36 ) | 1.71 ( 1.20 - 2.44 ) | 1.03 (0.73 - 1.45) |
| cholecalciferol | 256 / 281 | 447 / 788 | 1.50 ( 1.04 - 2.18 ) | 1.69 ( 1.26 - 2.27 ) | 1.76 (1.33 - 2.31) |
| azathioprine | 122 / 415 | 219 / 1016 | 1.42 ( 0.92 - 2.20 ) | 1.58 ( 1.11 - 2.25 ) | 1.23 (0.91 - 1.67) |
| tacrolimus | 124 / 413 | 197 / 1038 | 1.42 ( 0.88 - 2.27 ) | 1.56 ( 1.07 - 2.27 ) | 1.90 (1.40 - 2.58) |
| aspirin | 290 / 247 | 523 / 712 | 1.40 ( 0.96 - 2.05 ) | 1.55 ( 1.14 - 2.09 ) | 1.26 (0.96 - 1.66) |
| mycophenolate | 209 / 328 | 360 / 875 | 1.37 ( 0.95 - 1.98 ) | 1.50 ( 1.12 - 2.02 ) | 1.51 (1.15 - 1.99) |
| belimumab | 107 / 430 | 169 / 1066 | 1.29 ( 0.79 - 2.11 ) | 1.39 ( 0.94 - 2.06 ) | 2.06 (1.52 - 2.81) |
| levothyroxine | 166 / 371 | 298 / 937 | 1.21 ( 0.79 - 1.87 ) | 1.29 ( 0.91 - 1.82 ) | 0.91 (0.66 - 1.27) |
| estradiol | 129 / 408 | 212 / 1023 | 0.93 ( 0.58 - 1.47 ) | 0.91 ( 0.63 - 1.31 ) | 0.64 (0.43 - 0.96) |
| hydroxychloroquin<br>e | 433 / 104 | 1012 / 223 | 0.65 ( 0.41 - 1.03 ) | 0.58 ( 0.40 - 0.83 ) | 1.03 (0.66 - 1.61) |

**Supplementary Table 5.** Association between medication exposure and ADEs in rheumatic arthritis.

| Medication | Cases<br>(Exposed /<br>Unexposed ) | Controls<br>(Exposed /<br>Unexposed ) | Unadjusted OR | Adjusted OR<br>(demographics +<br>comorbidities) | Adjusted OR with<br>CRP (47% of<br>entire RA cohort) |
| --- | --- | --- | --- | --- | --- |
| acetaminophen | 988 / 158 | 1698 /<br>1163 | 4.67 ( 3.07 - 7.10 ) | 6.87 ( 4.99 - 9.45 ) | 3.11 ( 2.39 - 4.05 ) |
| lorazepam | 443 / 703 | 437 / 2424 | 4.07 ( 2.92 - 5.68 ) | 5.97 ( 4.61 - 7.72 ) | 2.81 ( 2.30 - 3.45 ) |
| famotidine | 482 / 664 | 533 / 2328 | 3.51 ( 2.58 - 4.78 ) | 4.94 ( 3.89 - 6.29 ) | 2.31 ( 1.90 - 2.81 ) |
| dexamethasone | 616 / 530 | 778 / 2083 | 3.40 ( 2.51 - 4.60 ) | 4.76 ( 3.77 - 6.03 ) | 2.34 ( 1.94 - 2.83 ) |
| pantoprazole | 554 / 592 | 609 / 2252 | 3.38 ( 2.50 - 4.59 ) | 4.71 ( 3.71 - 5.97 ) | 2.51 ( 2.07 - 3.05 ) |
| prednisone | 911 / 235 | 1718 /<br>1143 | 3.25 ( 2.31 - 4.57 ) | 4.50 ( 3.45 - 5.87 ) | 2.35 ( 1.83 - 3.00 ) |
| sennosides, USP | 463 / 683 | 445 / 2416 | 3.20 ( 2.34 - 4.38 ) | 4.43 ( 3.46 - 5.66 ) | 3.32 ( 2.73 - 4.05 ) |
| trazodone | 292 / 854 | 271 / 2590 | 3.08 ( 2.12 - 4.46 ) | 4.26 ( 3.18 - 5.71 ) | 2.64 ( 2.10 - 3.32 ) |
| oxycodone | 687 / 459 | 851 / 2010 | 2.92 ( 2.19 - 3.89 ) | 3.96 ( 3.16 - 4.96 ) | 2.56 ( 2.12 - 3.10 ) |
| hydromorphone | 484 / 662 | 510 / 2351 | 2.83 ( 2.09 - 3.82 ) | 3.80 ( 3.00 - 4.82 ) | 2.72 ( 2.24 - 3.31 ) |
| ketorolac | 438 / 708 | 459 / 2402 | 2.72 ( 2.01 - 3.68 ) | 3.65 ( 2.87 - 4.64 ) | 2.39 ( 1.96 - 2.92 ) |
| duloxetine | 302 / 844 | 364 / 2497 | 2.72 ( 1.96 - 3.77 ) | 3.64 ( 2.81 - 4.72 ) | 1.36 ( 1.07 - 1.73 ) |
| furosemide | 358 / 788 | 391 / 2470 | 2.70 ( 1.96 - 3.72 ) | 3.55 ( 2.75 - 4.57 ) | 2.31 ( 1.88 - 2.84 ) |
| budesonide | 280 / 866 | 316 / 2545 | 2.69 ( 1.90 - 3.80 ) | 3.55 ( 2.70 - 4.67 ) | 1.29 ( 1.01 - 1.66 ) |
| docusate | 484 / 662 | 646 / 2215 | 2.48 ( 1.85 - 3.33 ) | 3.22 ( 2.56 - 4.07 ) | 2.22 ( 1.83 - 2.69 ) |
| pregabalin | 228 / 918 | 272 / 2589 | 2.43 ( 1.72 - 3.44 ) | 3.11 ( 2.37 - 4.09 ) | 1.33 ( 1.02 - 1.73 ) |
| gabapentin | 683 / 463 | 1026 /<br>1835 | 2.41 ( 1.82 - 3.19 ) | 3.11 ( 2.49 - 3.88 ) | 1.76 ( 1.46 - 2.13 ) |
| methylprednisolone | 661 / 485 | 984 / 1877 | 2.40 ( 1.84 - 3.15 ) | 3.12 ( 2.52 - 3.86 ) | 2.04 ( 1.69 - 2.47 ) |
| omeprazole | 560 / 586 | 826 / 2035 | 2.27 ( 1.70 - 3.01 ) | 2.89 ( 2.30 - 3.63 ) | 1.91 ( 1.58 - 2.31 ) |
| amoxicillin | 580 / 566 | 833 / 2028 | 2.21 ( 1.68 - 2.91 ) | 2.79 ( 2.24 - 3.48 ) | 1.83 ( 1.51 - 2.21 ) |
| triamcinolone | 677 / 469 | 1122/1739 | 2.19 ( 1.67 - 2.86 ) | 2.77 ( 2.23 - 3.43 ) | 1.76 ( 1.45 - 2.13 ) |
| infliximab | 80 / 1066 | 93 / 2768 | 2.06 ( 1.20 - 3.55 ) | 2.55 ( 1.65 - 3.94 ) | 1.82 ( 1.23 - 2.69 ) |
| tramadol | 412 / 734 | 521 / 2340 | 2.03 ( 1.53 - 2.69 ) | 2.50 ( 1.99 - 3.14 ) | 1.87 ( 1.52 - 2.29 ) |
| fluticasone | 549 / 597 | 830 / 2031 | 1.91 ( 1.44 - 2.51 ) | 2.31 ( 1.85 - 2.89 ) | 1.73 ( 1.43 - 2.09 ) |
| albuterol | 532 / 614 | 798 / 2063 | 1.90 ( 1.46 - 2.48 ) | 2.31 ( 1.86 - 2.86 ) | 1.76 ( 1.46 - 2.13 ) |
| metformin | 204 / 942 | 281 / 2580 | 1.90 ( 1.35 - 2.68 ) | 2.32 ( 1.77 - 3.06 ) | 1.74 ( 1.37 - 2.23 ) |
| escitalopram | 200 / 946 | 255 / 2606 | 1.88 ( 1.28 - 2.75 ) | 2.29 ( 1.68 - 3.11 ) | 1.35 ( 1.02 - 1.79 ) |
| cyclobenzaprine | 386 / 760 | 508 / 2353 | 1.87 ( 1.38 - 2.52 ) | 2.23 ( 1.75 - 2.84 ) | 2.01 ( 1.65 - 2.46 ) |
| diclofenac | 606 / 540 | 1007/1854 | 1.82 ( 1.38 - 2.40 ) | 2.16 ( 1.73 - 2.70 ) | 1.40 ( 1.16 - 1.68 ) |
| bupropion | 179 / 967 | 250 / 2611 | 1.78 ( 1.23 - 2.56 ) | 2.13 ( 1.59 - 2.86 ) | 1.67 ( 1.28 - 2.19 ) |
| amlodipine | 382 / 764 | 629 / 2232 | 1.76 ( 1.32 - 2.37 ) | 2.09 ( 1.66 - 2.65 ) | 1.23 ( 1.00 - 1.51 ) |
| aspirin | 606 / 540 | 1033/1828 | 1.75 ( 1.33 - 2.29 ) | 2.05 ( 1.65 - 2.55 ) | 1.51 ( 1.25 - 1.83 ) |
| celecoxib | 460 / 686 | 817 / 2044 | 1.64 ( 1.25 - 2.16 ) | 1.91 ( 1.53 - 2.38 ) | 1.35 ( 1.12 - 1.64 ) |
| ibuprofen | 513 / 633 | 906 / 1955 | 1.55 ( 1.19 - 2.02 ) | 1.78 ( 1.44 - 2.20 ) | 1.71 ( 1.41 - 2.06 ) |
| atorvastatin | 408 / 738 | 792 / 2069 | 1.49 ( 1.13 - 1.96 ) | 1.69 ( 1.35 - 2.11 ) | 1.08 ( 0.89 - 1.32 ) |
| losartan | 298 / 848 | 474 / 2387 | 1.41 ( 1.03 - 1.93 ) | 1.57 ( 1.22 - 2.02 ) | 1.12 ( 0.89 - 1.40 ) |
| metoprolol | 378 / 768 | 572 / 2289 | 1.40 ( 1.04 - 1.86 ) | 1.55 ( 1.22 - 1.95 ) | 1.55 ( 1.26 - 1.91 ) |
| hydrochlorothiazide | 306 / 840 | 548 / 2313 | 1.37 ( 1.01 - 1.86 ) | 1.50 ( 1.17 - 1.92 ) | 1.35 ( 1.09 - 1.67 ) |
| levothyroxine | 365 / 781 | 684 / 2177 | 1.37 ( 1.02 - 1.83 ) | 1.50 ( 1.19 - 1.90 ) | 1.68 ( 1.38 - 2.05 ) |
| lisinopril | 232 / 914 | 382 / 2479 | 1.36 ( 0.96 - 1.92 ) | 1.49 ( 1.13 - 1.97 ) | 1.44 ( 1.13 - 1.82 ) |
| abatacept | 173 / 973 | 278 / 2583 | 1.33 ( 0.89 - 1.99 ) | 1.45 ( 1.05 - 2.01 ) | 1.43 ( 1.11 - 1.84 ) |
| rosuvastatin | 279 / 867 | 442 / 2419 | 1.29 ( 0.93 - 1.79 ) | 1.40 ( 1.07 - 1.82 ) | 1.43 ( 1.14 - 1.78 ) |
| tocilizumab | 135 / 1011 | 212 / 2649 | 1.24 ( 0.80 - 1.92 ) | 1.32 ( 0.93 - 1.90 ) | 1.70 ( 1.31 - 2.21 ) |
| estradiol | 285 / 861 | 565 / 2296 | 1.20 ( 0.88 - 1.62 ) | 1.27 ( 0.99 - 1.62 ) | 0.99 ( 0.79 - 1.24 ) |
| tofacitinib | 140 / 1006 | 243 / 2618 | 1.10 ( 0.73 - 1.67 ) | 1.14 ( 0.81 - 1.59 ) | 1.15 ( 0.87 - 1.52 ) |
| adalimumab | 259 / 887 | 555 / 2306 | 1.07 ( 0.78 - 1.48 ) | 1.09 ( 0.84 - 1.42 ) | 1.08 ( 0.87 - 1.34 ) |
| hydroxychloroquine | 606 / 540 | 1385/1476 | 0.92 ( 0.71 - 1.20 ) | 0.90 ( 0.72 - 1.11 ) | 1.11 ( 0.92 - 1.34 ) |
| methotrexate | 498 / 648 | 1289/1572 | 0.88 ( 0.67 - 1.13 ) | 0.84 ( 0.68 - 1.04 ) | 0.75 ( 0.62 - 0.90 ) |
| etanercept | 193 / 953 | 426 / 2435 | 0.84 ( 0.59 - 1.20 ) | 0.80 ( 0.60 - 1.06 ) | 1.19 ( 0.95 - 1.50 ) |
| sulfasalazine | 148 / 998 | 360 / 2501 | 0.78 ( 0.53 - 1.15 ) | 0.72 ( 0.53 - 0.99 ) | 1.01 ( 0.78 - 1.32 ) |

**Supplementary Table 6.** Association between medication exposure and ADEs in psoriatic arthritis.

| Medication | Cases<br>(Exposed<br>/<br>Unexposed) | Controls<br>(Exposed /<br>Unexposed) | Unadjusted OR | Adjusted OR<br>(demographics +<br>comorbidities) | Adjusted OR CRP<br>(36% of entire<br>PsA cohort) |
| --- | --- | --- | --- | --- | --- |
| acetaminophen | 443 / 89 | 688 / 648 | 6.21 ( 3.43- 11.24 ) | 9.67 ( 6.18- 15.14 ) | 2.47 ( 1.69 - 3.59 ) |
| pantoprazole | 242 / 290 | 230 / 1106 | 3.95 ( 2.59 - 6.02 ) | 5.78 ( 4.17 - 8.01 ) | 1.93 ( 1.39 - 2.69 ) |
| famotidine | 221 / 311 | 199 / 1137 | 3.67 ( 2.40 - 5.63 ) | 5.24 ( 3.76 - 7.30 ) | 2.55 ( 1.83 - 3.56 ) |
| hydromorphone | 256 / 276 | 255 / 1081 | 3.56 ( 2.34 - 5.43 ) | 5.05 ( 3.64 - 7.01 ) | 2.97 ( 2.17 - 4.08 ) |
| oxycodone | 317 / 215 | 380 / 956 | 3.45 ( 2.27 - 5.25 ) | 4.79 ( 3.46 - 6.63 ) | 2.43 ( 1.77 - 3.33 ) |
| furosemide | 156 / 376 | 143 / 1193 | 3.30 ( 2.09 - 5.21 ) | 4.63 ( 3.24 - 6.63 ) | 1.97 ( 1.35 - 2.88 ) |
| dexamethasone | 309 / 223 | 377 / 959 | 3.18 ( 2.11 - 4.80 ) | 4.37 ( 3.17 - 6.02 ) | 1.81 ( 1.33 - 2.48 ) |
| ketorolac | 211 / 321 | 243 / 1093 | 3.10 ( 1.96 - 4.91 ) | 4.21 ( 2.95 - 6.03 ) | 1.76 ( 1.26 - 2.44 ) |
| budesonide | 136 / 396 | 131 / 1205 | 3.01 ( 1.85 - 4.91 ) | 4.12 ( 2.81 - 6.05 ) | 1.08 ( 0.70 - 1.67 ) |
| sennosides, USP | 216 / 316 | 209 / 1127 | 2.98 ( 1.94 - 4.56 ) | 4.06 ( 2.90 - 5.67 ) | 2.59 ( 1.87 - 3.59 ) |
| bupropion | 96 / 436 | 131 / 1205 | 2.92 ( 1.69 - 5.07 ) | 3.95 ( 2.56 - 6.09 ) | 1.43 ( 0.94 - 2.17 ) |
| duloxetine | 137 / 395 | 135 / 1201 | 2.80 ( 1.73 - 4.52 ) | 3.76 ( 2.57 - 5.49 ) | 1.92 ( 1.31 - 2.80 ) |
| prednisone | 359 / 173 | 555 / 781 | 2.78 ( 1.82 - 4.25 ) | 3.69 ( 2.65 - 5.14 ) | 1.74 ( 1.24 - 2.42 ) |
| tramadol | 170 / 362 | 170 / 1166 | 2.76 ( 1.79 - 4.25 ) | 3.72 ( 2.64 - 5.24 ) | 1.72 ( 1.21 - 2.44 ) |
| gabapentin | 312 / 220 | 396 / 940 | 2.75 ( 1.85 - 4.10 ) | 3.68 ( 2.70 - 5.04 ) | 2.05 ( 1.50 - 2.80 ) |
| pregabalin | 100 / 432 | 99 / 1237 | 2.74 ( 1.62 - 4.64 ) | 3.70 ( 2.44 - 5.61 ) | 1.30 ( 0.82 - 2.04 ) |
| albuterol | 228 / 304 | 328 / 1008 | 2.64 ( 1.79 - 3.89 ) | 3.49 ( 2.57 - 4.74 ) | 1.49 ( 1.08 - 2.06 ) |
| cyclobenzaprine | 176 / 356 | 234 / 1102 | 2.50 ( 1.63 - 3.81 ) | 3.24 ( 2.32 - 4.53 ) | 1.31 ( 0.93 - 1.83 ) |
| cephalexin | 240 / 292 | 319 / 1017 | 2.48 ( 1.63 - 3.77 ) | 3.20 ( 2.30 - 4.45 ) | 1.37 ( 1.00 - 1.89 ) |
| azithromycin | 225 / 307 | 315 / 1021 | 2.40 ( 1.62 - 3.56 ) | 3.15 ( 2.30 - 4.30 ) | 1.52 ( 1.11 - 2.10 ) |
| omeprazole | 235 / 297 | 331 / 1005 | 2.36 ( 1.59 - 3.49 ) | 3.04 ( 2.23 - 4.14 ) | 1.46 ( 1.06 - 2.00 ) |
| hydrocortisone | 237 / 295 | 310 / 1026 | 2.28 ( 1.53 - 3.41 ) | 2.93 ( 2.13 - 4.04 ) | 2.73 ( 1.98 - 3.76 ) |
| triamcinolone | 378 / 154 | 697 / 639 | 2.24 ( 1.46 - 3.45 ) | 2.84 ( 2.01 - 3.99 ) | 1.67 ( 1.20 - 2.32 ) |
| infliximab | 46 / 486 | 60 / 1276 | 2.17 ( 1.12 - 4.21 ) | 2.76 ( 1.62 - 4.69 ) | 1.56 ( 0.91 - 2.67 ) |
| fluticasone | 236 / 296 | 336 / 1000 | 2.10 ( 1.40 - 3.16 ) | 2.61 ( 1.89 - 3.61 ) | 1.34 ( 0.97 - 1.85 ) |
| hydrochlorothiazide | 131 / 401 | 213 / 1123 | 2.08 ( 1.28 - 3.38 ) | 2.58 ( 1.76 - 3.80 ) | 1.55 ( 1.09 - 2.22 ) |
| aspirin | 251 / 281 | 413 / 923 | 2.07 ( 1.41 - 3.05 ) | 2.58 ( 1.89 - 3.51 ) | 0.92 ( 0.66 - 1.27 ) |
| methylprednisolone | 252 / 280 | 342 / 994 | 2.01 ( 1.38 - 2.95 ) | 2.49 ( 1.83 - 3.37 ) | 1.56 ( 1.14 - 2.13 ) |
| diclofenac | 250 / 282 | 421 / 915 | 2.01 ( 1.37 - 2.95 ) | 2.46 ( 1.81 - 3.34 ) | 1.00 ( 0.73 - 1.36 ) |
| celecoxib | 206 / 326 | 302 / 1034 | 2.00 ( 1.32 - 3.03 ) | 2.45 ( 1.76 - 3.41 ) | 1.54 ( 1.12 - 2.11 ) |
| amoxicillin | 239 / 293 | 344 / 992 | 1.92 ( 1.30 - 2.83 ) | 2.35 ( 1.72 - 3.20 ) | 1.45 ( 1.06 - 1.99 ) |
| atorvastatin | 202 / 330 | 347 / 989 | 1.90 ( 1.27 - 2.85 ) | 2.30 ( 1.67 - 3.17 ) | 1.03 ( 0.73 - 1.46 ) |
| metoprolol | 162 / 370 | 234 / 1102 | 1.87 ( 1.23 - 2.83 ) | 2.25 ( 1.61 - 3.13 ) | 1.12 ( 0.78 - 1.61 ) |
| clobetasol | 316 / 216 | 675 / 661 | 1.86 ( 1.25 - 2.76 ) | 2.24 ( 1.63 - 3.07 ) | 0.92 ( 0.68 - 1.26 ) |
| ustekinumab | 93 / 439 | 195 / 1141 | 1.82 ( 1.13 - 2.91 ) | 2.20 ( 1.50 - 3.22 ) | 1.12 ( 0.74 - 1.69 ) |
| ibuprofen | 237 / 295 | 437 / 899 | 1.73 ( 1.16 - 2.57 ) | 2.03 ( 1.48 - 2.78 ) | 1.54 ( 1.12 - 2.10 ) |
| leflunomide | 74 / 458 | 112 / 1224 | 1.63 ( 0.94 - 2.85 ) | 1.89 ( 1.21 - 2.95 ) | 1.49 ( 0.99 - 2.25 ) |
| lisinopril | 119 / 413 | 210 / 1126 | 1.57 ( 1.01 - 2.45 ) | 1.80 ( 1.26 - 2.58 ) | 1.30 ( 0.88 - 1.91 ) |
| metformin | 117 / 415 | 238 / 1098 | 1.57 ( 1.00 - 2.46 ) | 1.81 ( 1.26 - 2.59 ) | 0.72 ( 0.46 - 1.12 ) |
| fluocinonide | 198 / 334 | 377 / 959 | 1.53 ( 1.03 - 2.27 ) | 1.74 ( 1.27 - 2.40 ) | 1.29 ( 0.92 - 1.80 ) |
| hydroxychloroquin<br>e | 109 / 423 | 129 / 1207 | 1.48 ( 0.91 - 2.42 ) | 1.66 ( 1.12 - 2.47 ) | 1.75 ( 1.18 - 2.57 ) |
| levothyroxine | 133 / 399 | 216 / 1120 | 1.35 ( 0.89 - 2.06 ) | 1.50 ( 1.07 - 2.10 ) | 1.67 ( 1.16 - 2.38 ) |
| rosuvastatin | 116 / 416 | 213 / 1123 | 1.33 ( 0.83 - 2.12 ) | 1.44 ( 0.98 - 2.10 ) | 0.75 ( 0.49 - 1.15 ) |
| secukinumab | 137 / 395 | 312 / 1024 | 1.32 ( 0.87 - 2.01 ) | 1.44 ( 1.03 - 2.01 ) | 0.74 ( 0.52 - 1.07 ) |
| losartan | 120 / 412 | 202 / 1134 | 1.31 ( 0.84 - 2.03 ) | 1.42 ( 0.99 - 2.02 ) | 0.82 ( 0.52 - 1.30 ) |
| sulfasalazine | 114 / 418 | 218 / 1118 | 1.11 ( 0.69 - 1.77 ) | 1.14 ( 0.78 - 1.67 ) | 1.37 ( 0.97 - 1.94 ) |
| amlodipine | 143 / 389 | 267 / 1069 | 1.06 ( 0.71 - 1.58 ) | 1.08 ( 0.78 - 1.49 ) | 1.07 ( 0.75 - 1.53 ) |
| etanercept | 127 / 405 | 257 / 1079 | 0.92 ( 0.58 - 1.46 ) | 0.90 ( 0.62 - 1.31 ) | 1.21 ( 0.86 - 1.71 ) |
| adalimumab | 226 / 306 | 532 / 804 | 0.89 ( 0.61 - 1.29 ) | 0.85 ( 0.63 - 1.16 ) | 0.79 ( 0.58 - 1.08 ) |
| methotrexate | 233 / 299 | 562 / 774 | 0.75 ( 0.51 - 1.11 ) | 0.69 ( 0.51 - 0.95 ) | 0.90 ( 0.66 - 1.23 ) |

**Supplementary Table 7.** Association between medication exposure and ADEs in systemic sclerosis.

| Medication | Cases<br>(Exposed /<br>Unexposed ) | Controls<br>(Exposed /<br>Unexposed ) | Unadjusted OR | Adjusted OR<br>(demographics +<br>comorbidities) | Adjusted OR CRP<br>(30% of entire<br>SSc cohort) |
| --- | --- | --- | --- | --- | --- |
| rifaximin | 100 / 187 | 61 / 448 | 8.82 ( 4.03 - 19.33 ) | 14.41 ( 7.97 - 26.07 ) | 3.88 ( 2.34 - 6.43 ) |
| metoclopramide | 126 / 161 | 63 / 446 | 7.67 ( 3.63 - 16.21 ) | 12.32 ( 6.99 - 21.71 ) | 4.35 ( 2.67 - 7.09 ) |
| methylprednisolone | 129 / 158 | 69 / 440 | 5.63 ( 2.83 - 11.21 ) | 8.60 ( 5.08 - 14.56 ) | 4.61 ( 2.84 - 7.47 ) |
| magnesium sulfate | 136 / 151 | 67 / 442 | 5.60 ( 2.78 - 11.26 ) | 8.55 ( 5.00 - 14.63 ) | 3.50 ( 2.16 - 5.66 ) |
| enoxaparin | 127 / 160 | 69 / 440 | 5.28 ( 2.71 - 10.30 ) | 7.94 ( 4.75 - 13.26 ) | 4.51 ( 2.78 - 7.32 ) |
| azithromycin | 143 / 144 | 90 / 419 | 5.20 ( 2.58 - 10.45 ) | 7.68 ( 4.49 - 13.14 ) | 4.08 ( 2.52 - 6.60 ) |
| acetaminophen | 268 / 19 | 304 / 205 | 5.14 ( 1.98 - 13.33 ) | 7.21 ( 3.52 - 14.77 ) | 13.76 ( 3.32 - 56.96 ) |
| trimethoprim | 138 / 149 | 93 / 416 | 5.03 ( 2.64 - 9.61 ) | 7.38 ( 4.48 - 12.14 ) | 4.83 ( 2.97 - 7.85 ) |
| methadone | 19 / 268 | 9 / 500 | 4.70 ( 0.91 - 24.23 ) | 7.20 ( 2.04 - 25.37 ) | 9.38 ( 1.69 - 52.06 ) |
| hydromorphone | 167 / 120 | 110 / 399 | 3.86 ( 2.12 - 7.02 ) | 5.46 ( 3.43 - 8.68 ) | 4.24 ( 2.60 - 6.92 ) |
| morphine | 85 / 202 | 50 / 459 | 3.82 ( 1.92 - 7.59 ) | 5.37 ( 3.14 - 9.18 ) | 3.33 ( 1.99 - 5.58 ) |
| amoxicillin | 148 / 139 | 102 / 407 | 3.62 ( 1.97 - 6.68 ) | 4.98 ( 3.10 - 8.01 ) | 3.00 ( 1.87 - 4.81 ) |
| famotidine | 169 / 118 | 126 / 383 | 3.58 ( 1.94 - 6.60 ) | 4.92 ( 3.06 - 7.92 ) | 4.22 ( 2.60 - 6.86 ) |
| phenylephrine | 159 / 128 | 120 / 389 | 3.57 ( 1.91 - 6.67 ) | 4.92 ( 3.03 - 8.00 ) | 2.87 ( 1.78 - 4.64 ) |
| gabapentin | 182 / 105 | 163 / 346 | 3.54 ( 1.85 - 6.75 ) | 4.79 ( 2.91 - 7.88 ) | 2.58 ( 1.60 - 4.17 ) |
| methoxsalen | 7 / 280 | 3 / 506 | 3.37 ( 0.27 - 42.26 ) | 4.89 ( 0.68 - 35.19 ) | 0.00 ( 0.00 - Inf ) |
| furosemide | 148 / 139 | 101 / 408 | 3.35 ( 1.82 - 6.16 ) | 4.54 ( 2.82 - 7.31 ) | 3.76 ( 2.33 - 6.06 ) |
| doxycycline | 117 / 170 | 93 / 416 | 3.30 ( 1.81 - 6.03 ) | 4.48 ( 2.80 - 7.18 ) | 4.11 ( 2.54 - 6.65 ) |
| pantoprazole | 201 / 86 | 189 / 320 | 3.17 ( 1.71 - 5.90 ) | 4.12 ( 2.56 - 6.65 ) | 2.34 ( 1.42 - 3.85 ) |
| lorazepam | 137 / 150 | 81 / 428 | 3.07 ( 1.70 - 5.55 ) | 4.15 ( 2.61 - 6.61 ) | 4.60 ( 2.83 - 7.46 ) |
| metoprolol | 107 / 180 | 103 / 406 | 3.06 ( 1.54 - 6.10 ) | 4.06 ( 2.37 - 6.96 ) | 1.69 ( 1.04 - 2.76 ) |
| pregabalin | 57 / 230 | 36 / 473 | 2.95 ( 1.37 - 6.34 ) | 4.01 ( 2.20 - 7.34 ) | 2.92 ( 1.56 - 5.50 ) |
| cephalexin | 141 / 146 | 133 / 376 | 2.91 ( 1.63 - 5.20 ) | 3.76 ( 2.40 - 5.90 ) | 2.72 ( 1.69 - 4.37 ) |
| sennosides, USP | 158 / 129 | 113 / 396 | 2.80 ( 1.55 - 5.06 ) | 3.65 ( 2.30 - 5.79 ) | 3.35 ( 2.07 - 5.41 ) |
| prednisone | 188 / 99 | 201 / 308 | 2.71 ( 1.51 - 4.88 ) | 3.47 ( 2.19 - 5.49 ) | 4.22 ( 2.42 - 7.34 ) |
| albuterol | 153 / 134 | 158 / 351 | 2.71 ( 1.51 - 4.86 ) | 3.45 ( 2.19 - 5.45 ) | 1.53 ( 0.96 - 2.44 ) |
| tacrolimus | 63 / 224 | 52 / 457 | 2.55 ( 1.27 - 5.10 ) | 3.29 ( 1.90 - 5.70 ) | 3.53 ( 1.99 - 6.29 ) |
| acyclovir | 56 / 231 | 34 / 475 | 2.43 ( 1.10 - 5.33 ) | 3.08 ( 1.65 - 5.72 ) | 3.79 ( 2.07 - 6.93 ) |
| esomeprazole | 81 / 206 | 71 / 438 | 2.42 ( 1.31 - 4.48 ) | 3.07 ( 1.88 - 5.00 ) | 3.08 ( 1.84 - 5.14 ) |
| oxycodone | 194 / 93 | 165 / 344 | 2.38 ( 1.37 - 4.13 ) | 2.97 ( 1.92 - 4.58 ) | 3.07 ( 1.85 - 5.11 ) |
| dexamethasone | 181 / 106 | 149 / 360 | 2.34 ( 1.30 - 4.19 ) | 2.87 ( 1.82 - 4.54 ) | 1.75 ( 1.10 - 2.79 ) |
| zolpidem | 81 / 206 | 73 / 436 | 2.33 ( 1.22 - 4.42 ) | 2.98 ( 1.79 - 4.96 ) | 1.42 ( 0.81 - 2.49 ) |
| lisinopril | 70 / 217 | 84 / 425 | 2.10 ( 1.07 - 4.13 ) | 2.57 ( 1.50 - 4.40 ) | 2.28 ( 1.32 - 3.94 ) |
| docusate | 145 / 142 | 128 / 381 | 2.04 ( 1.16 - 3.59 ) | 2.45 ( 1.57 - 3.83 ) | 2.89 ( 1.80 - 4.64 ) |
| triamcinolone | 165 / 122 | 192 / 317 | 2.03 ( 1.19 - 3.47 ) | 2.45 ( 1.61 - 3.74 ) | 1.53 ( 0.96 - 2.45 ) |
| hydroxychloroquin<br>e | 131 / 156 | 198 / 311 | 2.00 ( 1.10 - 3.65 ) | 2.37 ( 1.48 - 3.79 ) | 1.26 ( 0.79 - 2.01 ) |
| sildenafil | 126 / 161 | 191 / 318 | 1.65 ( 0.93 - 2.94 ) | 1.88 ( 1.19 - 2.96 ) | 1.55 ( 0.97 - 2.46 ) |
| spironolactone | 59 / 228 | 44 / 465 | 1.64 ( 0.80 - 3.36 ) | 1.88 ( 1.07 - 3.32 ) | 2.11 ( 1.20 - 3.70 ) |
| fluticasone | 131 / 156 | 145 / 364 | 1.60 ( 0.88 - 2.92 ) | 1.78 ( 1.11 - 2.86 ) | 1.35 ( 0.85 - 2.14 ) |
| aspirin | 177 / 110 | 205 / 304 | 1.53 ( 0.88 - 2.67 ) | 1.71 ( 1.10 - 2.64 ) | 2.18 ( 1.35 - 3.54 ) |
| omeprazole | 185 / 102 | 267 / 242 | 1.49 ( 0.86 - 2.58 ) | 1.65 ( 1.07 - 2.55 ) | 1.13 ( 0.69 - 1.84 ) |
| amlodipine | 139 / 148 | 201 / 308 | 1.47 ( 0.84 - 2.57 ) | 1.63 ( 1.04 - 2.54 ) | 2.03 ( 1.26 - 3.25 ) |
| atorvastatin | 101 / 186 | 104 / 405 | 1.47 ( 0.80 - 2.69 ) | 1.63 ( 1.01 - 2.63 ) | 2.19 ( 1.33 - 3.60 ) |
| duloxetine | 54 / 233 | 46 / 463 | 1.42 ( 0.70 - 2.90 ) | 1.56 ( 0.89 - 2.75 ) | 1.41 ( 0.76 - 2.60 ) |
| losartan | 63 / 224 | 61 / 448 | 1.40 ( 0.71 - 2.77 ) | 1.54 ( 0.89 - 2.68 ) | 0.96 ( 0.49 - 1.88 ) |
| nifedipine | 83 / 204 | 130 / 379 | 1.25 ( 0.70 - 2.23 ) | 1.33 ( 0.84 - 2.10 ) | 0.80 ( 0.47 - 1.37 ) |
| escitalopram | 72 / 215 | 109 / 400 | 1.15 ( 0.62 - 2.14 ) | 1.19 ( 0.73 - 1.95 ) | 1.01 ( 0.61 - 1.68 ) |
| methotrexate | 69 / 218 | 89 / 420 | 0.98 ( 0.48 - 2.00 ) | 0.97 ( 0.55 - 1.72 ) | 1.68 ( 1.01 - 2.80 ) |
| rosuvastatin | 51 / 236 | 63 / 446 | 0.81 ( 0.36 - 1.84 ) | 0.77 ( 0.40 - 1.47 ) | 0.89 ( 0.43 - 1.83 ) |
| levothyroxine | 85 / 202 | 135 / 374 | 0.81 ( 0.46 - 1.44 ) | 0.77 ( 0.49 - 1.21 ) | 1.45 ( 0.89 - 2.36 ) |

**Supplementary Table 8.**
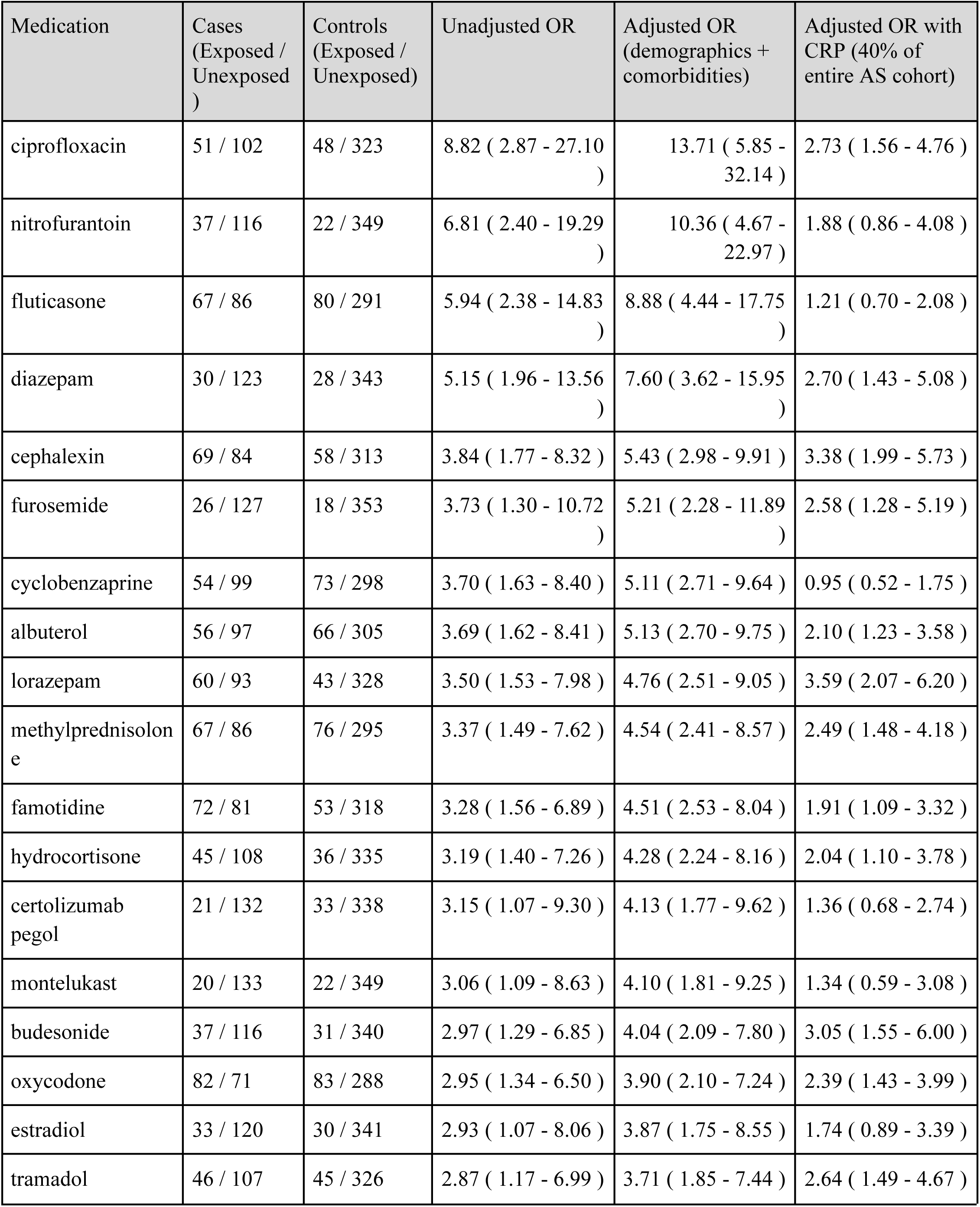
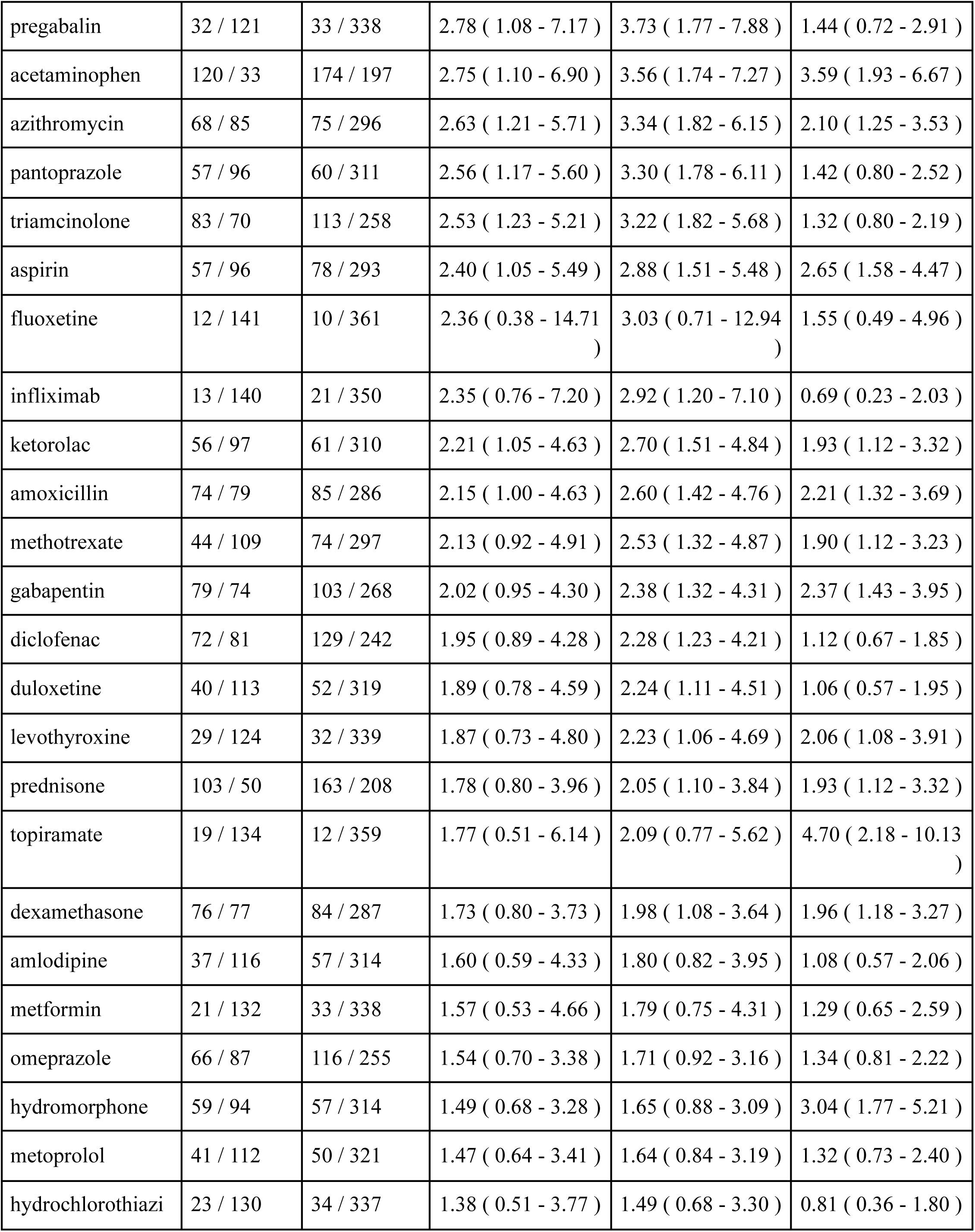
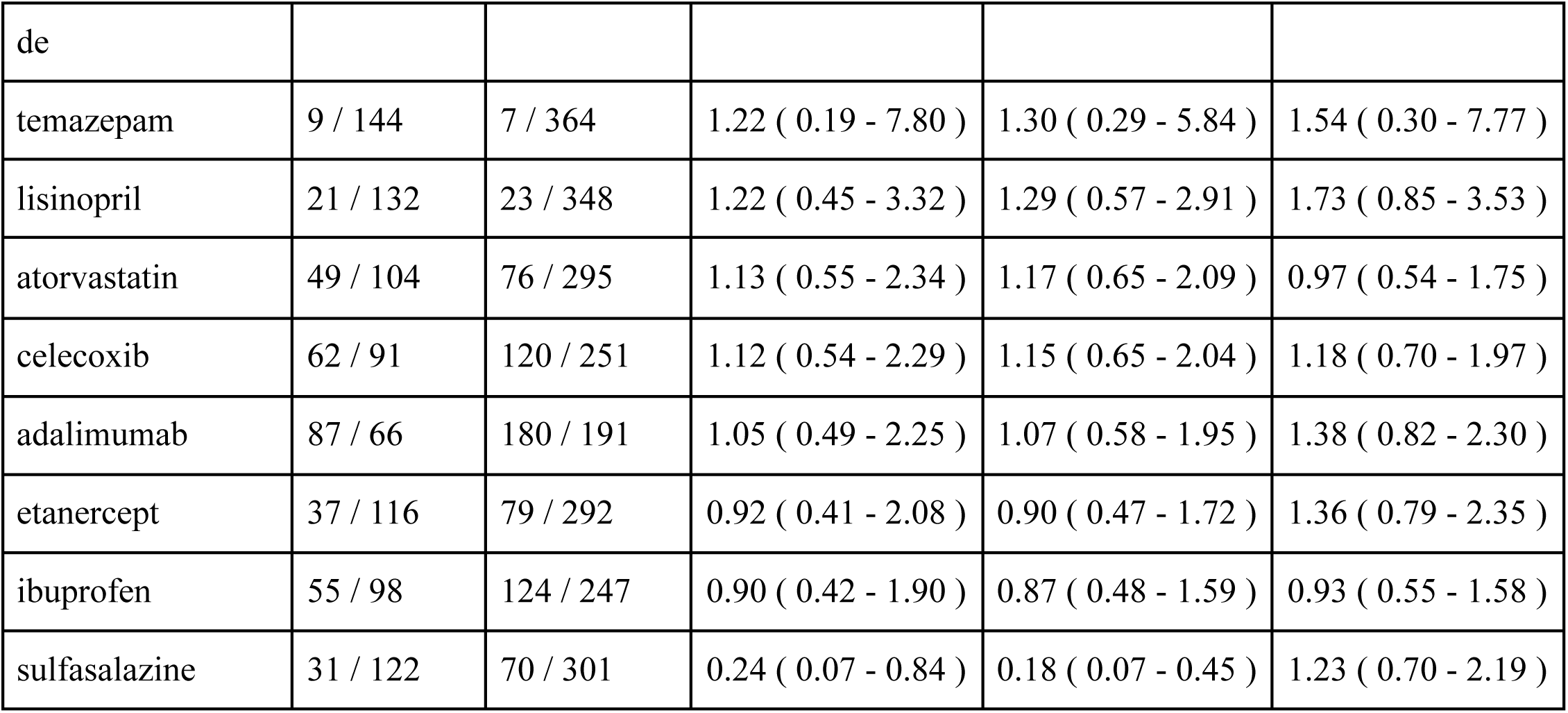
Association between medication exposure and ADEs in ankylosing spondylitis.

**Supplementary Table 9.** Association between medication exposure and ADEs in Sjögren’s disease.

| Medication | Cases<br>(Exposed /<br>Unexposed ) | Controls<br>(Exposed /<br>Unexposed) | Unadjusted OR | Adjusted OR<br>(demographics +<br>comorbidities) | Adjusted OR with<br>ESR (36% of<br>entire SjD cohort) |
| --- | --- | --- | --- | --- | --- |
| ipratropium | 147 / 352 | 124 / 971 | 4.07 ( 2.40 - 6.91 ) | 5.88 ( 3.89 - 8.87 ) | 1.95 ( 1.34 - 2.85 ) |
| famotidine | 210 / 289 | 212 / 883 | 4.04 ( 2.48 - 6.56 ) | 5.78 ( 3.96 - 8.41 ) | 2.05 ( 1.46 - 2.87 ) |
| trazodone | 117 / 382 | 110 / 985 | 3.97 ( 2.22 - 7.11 ) | 5.82 ( 3.70 - 9.15 ) | 1.65 ( 1.06 - 2.56 ) |
| acetaminophen | 417 / 82 | 604 / 491 | 3.36 ( 1.82 - 6.20 ) | 4.52 ( 2.82 - 7.24 ) | 3.58 ( 2.25 - 5.71 ) |
| lorazepam | 213 / 286 | 217 / 878 | 3.14 ( 1.95 - 5.03 ) | 4.24 ( 2.93 - 6.14 ) | 2.52 ( 1.80 - 3.53 ) |
| pantoprazole | 221 / 278 | 240 / 855 | 2.94 ( 1.87 - 4.63 ) | 3.93 ( 2.75 - 5.61 ) | 1.76 ( 1.26 - 2.45 ) |
| clonazepam | 66 / 433 | 70 / 1025 | 2.93 ( 1.38 - 6.22 ) | 3.90 ( 2.15 - 7.05 ) | 1.56 ( 0.87 - 2.76 ) |
| hydromorphone | 193 / 306 | 205 / 890 | 2.87 ( 1.80 - 4.58 ) | 3.83 ( 2.65 - 5.53 ) | 1.91 ( 1.36 - 2.68 ) |
| methylprednisolone | 252 / 247 | 299 / 796 | 2.78 ( 1.78 - 4.33 ) | 3.69 ( 2.60 - 5.23 ) | 1.61 ( 1.16 - 2.23 ) |
| oxycodone | 280 / 219 | 323 / 772 | 2.75 ( 1.76 - 4.31 ) | 3.60 ( 2.53 - 5.12 ) | 1.69 ( 1.22 - 2.35 ) |
| pregabalin | 114 / 385 | 104 / 991 | 2.59 ( 1.46 - 4.60 ) | 3.37 ( 2.14 - 5.31 ) | 2.11 ( 1.39 - 3.20 ) |
| triamcinolone | 329 / 170 | 427 / 668 | 2.56 ( 1.63 - 4.01 ) | 3.28 ( 2.30 - 4.66 ) | 1.58 ( 1.13 - 2.21 ) |
| budesonide | 140 / 359 | 144 / 951 | 2.54 ( 1.54 - 4.18 ) | 3.27 ( 2.20 - 4.86 ) | 1.68 ( 1.15 - 2.47 ) |
| albuterol | 253 / 246 | 328 / 767 | 2.52 ( 1.61 - 3.95 ) | 3.22 ( 2.26 - 4.58 ) | 1.81 ( 1.30 - 2.51 ) |
| omeprazole | 251 / 248 | 342 / 753 | 2.44 ( 1.57 - 3.80 ) | 3.09 ( 2.18 - 4.38 ) | 1.37 ( 0.99 - 1.90 ) |
| furosemide | 120 / 379 | 113 / 982 | 2.43 ( 1.50 - 3.95 ) | 3.13 ( 2.14 - 4.59 ) | 1.97 ( 1.31 - 2.96 ) |
| duloxetine | 127 / 372 | 153 / 942 | 2.30 ( 1.40 - 3.76 ) | 2.96 ( 2.00 - 4.39 ) | 1.44 ( 0.96 - 2.15 ) |
| prednisone | 348 / 151 | 510 / 585 | 2.29 ( 1.43 - 3.67 ) | 2.83 ( 1.96 - 4.08 ) | 1.69 ( 1.18 - 2.43 ) |
| gabapentin | 283 / 216 | 408 / 687 | 2.29 ( 1.48 - 3.53 ) | 2.87 ( 2.03 - 4.05 ) | 1.31 ( 0.95 - 1.82 ) |
| cephalexin | 227 / 272 | 283 / 812 | 2.25 ( 1.45 - 3.50 ) | 2.84 ( 2.01 - 4.02 ) | 1.64 ( 1.18 - 2.29 ) |
| dexamethasone | 287 / 212 | 339 / 756 | 2.19 ( 1.41 - 3.40 ) | 2.71 ( 1.91 - 3.82 ) | 1.79 ( 1.29 - 2.49 ) |
| bupropion | 87 / 412 | 105 / 990 | 2.18 ( 1.21 - 3.93 ) | 2.70 ( 1.69 - 4.30 ) | 1.89 ( 1.23 - 2.92 ) |
| clobetasol | 166 / 333 | 198 / 897 | 2.11 ( 1.35 - 3.30 ) | 2.60 ( 1.83 - 3.71 ) | 1.08 ( 0.74 - 1.57 ) |
| cyclobenzaprine | 180 / 319 | 208 / 887 | 2.11 ( 1.30 - 3.40 ) | 2.57 ( 1.76 - 3.76 ) | 1.42 ( 1.00 - 2.02 ) |
| sertraline | 93 / 406 | 101 / 994 | 2.09 ( 1.18 - 3.72 ) | 2.59 ( 1.64 - 4.10 ) | 1.11 ( 0.67 - 1.85 ) |
| hydrocortisone | 176 / 323 | 177 / 918 | 2.07 ( 1.32 - 3.24 ) | 2.54 ( 1.77 - 3.63 ) | 2.00 ( 1.40 - 2.84 ) |
| montelukast | 108 / 391 | 146 / 949 | 2.02 ( 1.19 - 3.42 ) | 2.47 ( 1.62 - 3.76 ) | 2.22 ( 1.51 - 3.25 ) |
| fluconazole | 146 / 353 | 159 / 936 | 1.92 ( 1.16 - 3.17 ) | 2.30 ( 1.54 - 3.42 ) | 1.90 ( 1.32 - 2.75 ) |
| celecoxib | 163 / 336 | 238 / 857 | 1.89 ( 1.19 - 3.00 ) | 2.25 ( 1.56 - 3.24 ) | 0.83 ( 0.57 - 1.23 ) |
| fluticasone | 277 / 222 | 374 / 721 | 1.87 ( 1.23 - 2.86 ) | 2.22 ( 1.59 - 3.10 ) | 1.43 ( 1.03 - 1.99 ) |
| nitrofurantoin | 176 / 323 | 203 / 892 | 1.85 ( 1.18 - 2.91 ) | 2.21 ( 1.54 - 3.17 ) | 1.19 ( 0.82 - 1.74 ) |
| azithromycin | 243 / 256 | 318 / 777 | 1.85 ( 1.22 - 2.81 ) | 2.21 ( 1.59 - 3.08 ) | 1.77 ( 1.27 - 2.45 ) |
| estradiol | 218 / 281 | 327 / 768 | 1.82 ( 1.17 - 2.85 ) | 2.17 ( 1.52 - 3.10 ) | 1.26 ( 0.90 - 1.76 ) |
| ketorolac | 182 / 317 | 206 / 889 | 1.82 ( 1.15 - 2.89 ) | 2.13 ( 1.48 - 3.08 ) | 1.36 ( 0.96 - 1.94 ) |
| amoxicillin | 269 / 230 | 347 / 748 | 1.80 ( 1.17 - 2.76 ) | 2.12 ( 1.51 - 2.98 ) | 1.47 ( 1.06 - 2.03 ) |
| losartan | 115 / 384 | 153 / 942 | 1.63 ( 0.99 - 2.71 ) | 1.89 ( 1.26 - 2.83 ) | 0.56 ( 0.33 - 0.93 ) |
| valacyclovir | 138 / 361 | 170 / 925 | 1.52 ( 0.92 - 2.52 ) | 1.70 ( 1.14 - 2.55 ) | 2.30 ( 1.61 - 3.28 ) |
| ibuprofen | 248 / 251 | 383 / 712 | 1.44 ( 0.95 - 2.20 ) | 1.60 ( 1.14 - 2.24 ) | 1.69 ( 1.22 - 2.34 ) |
| tramadol | 159 / 340 | 171 / 924 | 1.43 ( 0.89 - 2.29 ) | 1.58 ( 1.08 - 2.30 ) | 1.18 ( 0.80 - 1.72 ) |
| levothyroxine | 211 / 288 | 342 / 753 | 1.42 ( 0.91 - 2.22 ) | 1.58 ( 1.10 - 2.26 ) | 1.69 ( 1.21 - 2.36 ) |
| diclofenac | 253 / 246 | 347 / 748 | 1.39 ( 0.92 - 2.12 ) | 1.53 ( 1.10 - 2.15 ) | 0.98 ( 0.71 - 1.37 ) |
| amlodipine | 138 / 361 | 204 / 891 | 1.37 ( 0.86 - 2.16 ) | 1.50 ( 1.04 - 2.17 ) | 0.89 ( 0.59 - 1.33 ) |
| hydroxychloroquine | 276 / 223 | 586 / 509 | 1.28 ( 0.85 - 1.94 ) | 1.38 ( 0.99 - 1.93 ) | 1.06 ( 0.76 - 1.48 ) |
| methotrexate | 113 / 386 | 196 / 899 | 1.27 ( 0.74 - 2.18 ) | 1.34 ( 0.88 - 2.07 ) | 0.91 ( 0.63 - 1.32 ) |
| lisinopril | 79 / 420 | 104 / 991 | 1.22 ( 0.69 - 2.15 ) | 1.29 ( 0.82 - 2.04 ) | 1.49 ( 0.95 - 2.33 ) |
| abatacept | 39 / 460 | 57 / 1038 | 1.18 ( 0.45 - 3.09 ) | 1.22 ( 0.58 - 2.59 ) | 1.28 ( 0.75 - 2.19 ) |
| metoprolol | 147 / 352 | 206 / 889 | 1.17 ( 0.73 - 1.87 ) | 1.22 ( 0.84 - 1.78 ) | 1.39 ( 0.98 - 1.98 ) |
| aspirin | 238 / 261 | 359 / 736 | 1.12 ( 0.74 - 1.70 ) | 1.15 ( 0.83 - 1.61 ) | 1.29 ( 0.93 - 1.79 ) |
| atorvastatin | 148 / 351 | 244 / 851 | 1.11 ( 0.72 - 1.72 ) | 1.15 ( 0.81 - 1.63 ) | 0.87 ( 0.60 - 1.26 ) |
| hydrochlorothiazide | 94 / 405 | 156 / 939 | 0.80 ( 0.46 - 1.40 ) | 0.76 ( 0.49 - 1.18 ) | 0.84 ( 0.54 - 1.30 ) |

**Supplementary Table 10.** Prevalence of top 30 ADE-related ICD-10 codes by condition*.

| ICD10 | Name | SLE | RA | PsA | SSc | AS | SjD |
| --- | --- | --- | --- | --- | --- | --- | --- |
| K52.9 | Noninfective gastroenteritis and colitis, unspecified | 22.91% | 22.34% | 27.07% | 31.36% | 34.64% | 23.05% |
| F11.20 | Opioid dependence, uncomplicated | 4.84% | 8.12% | 4.89% | 2.09% | 9.15% | 4.01% |
| F11.90 | Opioid use, unspecified, uncomplicated | 2.23% | 5.76% | 4.89% | 2.44% | 3.27% | 2.20% |
| T78.40XA | Allergy, unspecified, initial encounter | 5.77% | 5.41% | 5.45% | 3.83% | 7.84% | 6.41% |
| L27.0 | Generalized skin eruption due to drugs and medicaments | 4.28% | 3.05% | 2.44% | 4.88% | 1.96% | 3.61% |
| K25.9 | Gastric ulcer, Unspecified as acute or chronic, without haemorrhage or perforation | 2.42% | 2.88% | 3.01% | 5.23% | 3.27% | 3.41% |
| K27.9 | Peptic ulcer, site unspecified, unspecified as acute or chronic, without haemorrhage or perforation | 2.42% | 2.44% | 2.63% | 2.09% | 0.65% | 3.01% |
| T46.991A | Poisoning by other agents primarily affecting the cardiovascular system, accidental (unintentional), initial encounter | 2.05% | 2.44% | 2.26% | 4.53% | 1.31% | 2.81% |
| E03.2 | Hypothyroidism due to medicaments and other exogenous substances | 0.74% | 2.36% | 1.69% | 0.35% | 0.00% | 2.20% |
| K29.00 | Acute gastritis without bleeding | 1.86% | 2.27% | 3.01% | 1.39% | 1.96% | 2.40% |
| G44.40 | Drug-induced headache, not elsewhere classified, not intractable | 2.42% | 2.01% | 3.20% | 1.74% | 1.31% | 3.21% |
| A04.72 | Enterocolitis due to Clostridium difficile, not specified as recurrent | 1.30% | 1.75% | 1.32% | 2.09% | 2.61% | 2.00% |
| I95.2 | Hypotension due to drugs | 0.37% | 1.48% | 0.94% | 3.48% | 0.65% | 1.20% |
| G62.0 | Drug-induced polyneuropathy | 0.56% | 1.40% | 1.32% | 1.74% | 0.65% | 1.40% |
| T45.1X5S | Adverse effect of antineoplastic and immunosuppressive drugs, sequela | 0.19% | 1.40% | 0.75% | 1.05% | 2.61% | 0.60% |
| T78.40XD | Allergy, unspecified, subsequent encounter | 0.74% | 1.40% | 1.88% | 0.70% | 0.00% | 2.00% |
| T78.3XXA | Angioneurotic edema, initial encounter | 2.42% | 1.31% | 1.50% | 1.05% | 0.65% | 3.01% |
| K25.4 | Gastric ulcer, Chronic or unspecified with haemorrhage | 0.56% | 1.22% | 0.56% | 1.05% | 0.65% | 0.60% |
| K52.1 | Toxic gastroenteritis and colitis | 1.12% | 1.22% | 1.13% | 2.09% | 1.96% | 1.60% |
| G72.0 | Drug-induced myopathy | 0.74% | 1.05% | 0.38% | 0.00% | 1.31% | 1.20% |
| K26.9 | Duodenal ulcer, unspecified as acute or chronic, without haemorrhage or perforation | 1.12% | 0.96% | 1.50% | 2.79% | 2.61% | 0.80% |
| T38.0X5S | Adverse effect of glucocorticoids and synthetic analogues, sequela | 1.12% | 0.96% | 0.00% | 0.00% | 0.00% | 0.00% |
| T78.40XS | Allergy, unspecified, sequela | 0.93% | 0.96% | 2.07% | 0.00% | 0.65% | 1.40% |
| T80.90XA | Unspecified complication following infusion and therapeutic injection, initial encounter | 0.93% | 0.96% | 1.50% | 1.05% | 0.00% | 0.60% |
| A04.7 | Enterocolitis due to Clostridium difficile | 1.30% | 0.09% | 0.19% | 1.74% | 0.00% | 0.40% |
| T50.901A | Poisoning by unspecified drugs, medicaments and biological substances, accidental (unintentional), initial encounter | 1.12% | 0.87% | 1.32% | 0.00% | 1.96% | 0.00% |
| K71.9 | Toxic liver disease, unspecified | 1.30% | 0.79% | 1.69% | 1.74% | 1.96% | 0.80% |
| L27.1 | Localized skin eruption due to drugs and medicaments | 1.30% | 0.79% | 0.56% | 0.35% | 1.31% | 1.20% |
| E27.3 | Drug-induced adrenocortical insufficiency | 0.56% | 0.70% | 0.38% | 0.00% | 0.00% | 0.00% |
| T78.2XXA | Anaphylactic shock, unspecified, initial encounter | 0.37% | 0.70% | 0.75% | 0.00% | 0.65% | 1.40% |

\*Reported ICD-10 codes reflect the top 30 codes observed among RA cases. Overlap of these top codes with other conditions was: SLE (80%), PsA (86.7%), SSc (70%), AS (67%), and SjD (80%).

## Notes

**Funding** LRA, NIAMS

### Competing Interest Statement

The authors have declared no competing interest.

### Funding Statement

This study was funded by LRA and NIAMS.

### Author Declarations

The Institutional Review Board of Stanford University gave ethical approval for this work.

## Reference

1 Abend AH, He I, Bahroos N, et al. Estimation of prevalence of autoimmune diseases in the United States using electronic health record data. J Clin Invest 2024; 135. DOI:10.1172/JCI178722.

2 Liu E, Perl A. Pathogenesis and treatment of autoimmune rheumatic diseases. Curr Opin Rheumatol 2019; 31: 307–15.

3 Grange L, Guilpain P, Truchetet M-E, Cracowski J-L, French Society of Pharmacology and Therapeutics. Challenges of autoimmune rheumatic disease treatment during the COVID-19 pandemic: A review. Therapie 2020; 75: 335–42.

4 Falasinnu T, Chaichian Y, Li J, et al. Does SLE widen or narrow race/ethnic disparities in the risk of five co-morbid conditions? Evidence from a community-based outpatient care system. Lupus 2019; 28: 1619–27.

5 Falasinnu T, Nguyen T, Jiang TE, et al. The Problem of Pain in Rheumatology: Clinical Profiles Associated With Concomitant Diagnoses With Chronic Overlapping Pain Conditions. ACR Open Rheumatol 2022; 4: 890–6.

6 Lewis J, Östör AJK. The prevalence and impact of polypharmacy in rheumatology. Rheumatology (Oxford) 2023; 62: SI237–41.

7 Krstić N, Stefanović N, Petronijević M, Damnjanović I. Polypharmacy and the risk of drug-drug interactions in patients with rheumatoid arthritis. Acta Fac Medicae Naissensis 2024; 41: 223–33.

8 Boukhlal S, Chouchana L, Saadi M, et al. Polypharmacy, drug-drug interactions, and adverse drug reactions among systemic sclerosis patients: A cross-sectional risk factor study. Semin Arthritis Rheum 2024; 67: 152469.

9 Miyake H, Sada RM, Akebo H, Tsugihashi Y, Hatta K. Polypharmacy prevalence and associated factors in patients with systemic lupus erythematosus: A single-centre, cross-sectional study. Mod Rheumatol 2023; 34: 106–12.

10 Berthelot W, Sirois C, Julien A-S, et al. The association between polypharmacy and disease control in rheumatoid arthritis and systemic lupus erythematosus: a cohort study. Rheumatol Int 2025; 45: 44.

11 Boeing LB, Fogaça NS, Kahlow BS, Skare T, Nisihara R. Polypharmacy and drug interactions in the management of rheumatoid arthritis. Rev Assoc Med Bras 2025; 71: e20250151.

12 Stanciu M, Lee J-YE, McDonald EG, et al. Medication-related hospitalisations in patients with SLE. Lupus Sci Med 2025; 12. DOI:10.1136/lupus-2024-001362.

13 Falasinnu T, Lu D, Baker MC. Annual trends in pain management modalities in patients with newly diagnosed autoimmune rheumatic diseases in the USA from 2007 to 2021: an administrative claims-based study. Lancet Rheumatol 2024; 6: e518–27.

14 Hohl CM, Karpov A, Reddekopp L, Doyle-Waters M, Stausberg J. ICD-10 codes used to identify adverse drug events in administrative data: a systematic review. J Am Med Inform Assoc 2014; 21: 547–57.

15 Juhásová Z, Karapinar-Çarkit F, Weir DL. The use of international classification of diseases codes to identify hospital admissions linked with adverse drug events: Validation study. Br J Clin Pharmacol 2025; 91: 2910–8.

16 WHOCC. ATCDDD - ATC/DDD Index. https://atcddd.fhi.no/atc_ddd_index/ (accessed Feb 27, 2026).

17 Charlson ME, Pompei P, Ales KL, MacKenzie CR. A new method of classifying prognostic comorbidity in longitudinal studies: development and validation. J Chronic Dis 1987; 40: 373–83.

18 Boukhlal S, Chouchana L, Saadi M, et al. Polypharmacy, drug-drug interactions, and adverse drug reactions among systemic sclerosis patients: A cross-sectional risk factor study. Semin Arthritis Rheum 2024; 67: 152469.

19 Filkova M, Carvalho J, Norton S, et al. Polypharmacy and Unplanned Hospitalizations in Patients with Rheumatoid Arthritis. J Rheumatol 2017; 44: 1786–93.

20 Smolen JS, Landewé RBM, Bergstra SA, et al. EULAR recommendations for the management of rheumatoid arthritis with synthetic and biological disease-modifying antirheumatic drugs: 2022 update. Ann Rheum Dis 2023; 82: 3–18.

21 Vergne-Salle P, Pouplin S, Trouvin AP, et al. The burden of pain in rheumatoid arthritis: Impact of disease activity and psychological factors. Eur J Pain 2020; 24: 1979–89.

22 Le N, Kenney MO, Walker A, et al. Visualizing real-world pain treatment pathways in chronic disease: A sequence-based analysis of polypharmacy in systemic lupus erythematosus. ACR Open Rheumatol 2025; 7: e70116.

23 Center for Drug Evaluation, Research. FDA Drug Safety Communication: FDA warns about serious risks and death when combining opioid pain or cough medicines with benzodiazepines; requires its strongest warning. U.S. Food and Drug Administration. 2024; published online Aug 26. https://www.fda.gov/drugs/drug-safety-and-availability/fda-drug-safety-communication-fda-warns-about-serious-risks-and-death-when-combining-opioid-pain-or (accessed Feb 2, 2026).

24 Filkova M, Carvalho J, Norton S, et al. Polypharmacy and Unplanned Hospitalizations in Patients with Rheumatoid Arthritis. J Rheumatol 2017; 44: 1786–93.

25 McMahan ZH, Kulkarni S, Chen J, et al. Systemic sclerosis gastrointestinal dysmotility: risk factors, pathophysiology, diagnosis and management. Nat Rev Rheumatol 2023; 19: 166–81.

26 Frede N, Rieger E, Lorenzetti R, et al. Respiratory tract infections and risk factors for infection in a cohort of 330 patients with axial spondyloarthritis or psoriatic arthritis. Front Immunol 2022; 13: 1040725.

27 Fauthoux T, Brisou D, Lazaro E, et al. Comorbidity burden on mortality in patients with systemic sclerosis. RMD Open 2024; 10: e004637.

28 Alex G, Shanoj KC, Varghese DR, Sageer Babu AS, Reji R, Shenoy PD. Co prescription of anti-acid therapy reduces the bioavailability of mycophenolate mofetil in systemic sclerosis patients: A crossover trial. Semin Arthritis Rheum 2023; 63: 152270.

29 Narum S, Westergren T, Klemp M. Corticosteroids and risk of gastrointestinal bleeding: a systematic review and meta-analysis. BMJ Open 2014; 4: e004587.

30 Acton EK, Willis AW, Hennessy S. Core concepts in pharmacoepidemiology: Key biases arising in pharmacoepidemiologic studies. Pharmacoepidemiol Drug Saf 2023; 32: 9–18.

